# Cooperative Architecture of Mitochondrial Proteome Homeostasis

**DOI:** 10.64898/2026.02.06.26345691

**Authors:** Patrick Forny, Merima Forny, Andrew J. Smith, Andrew Y. Sung, Kaixian Liu, David J. Pagliarini

## Abstract

Mitochondria are semi-autonomous organelles whose generation and maintenance demand precise expression, processing, and assembly of >1,000 proteins encoded across two genomes. To explore this cooperativity, we performed multiomic analyses on >200 cell lines harboring mitochondrial gene perturbations, generating >26M molecular measurements. Our data reveal that mitochondrial proteome homeostasis is heavily influenced by post-transcriptional processes. Through nearest neighbor analyses, we reveal diverse protein activities undergirding this regulation, including MDH2’s regulation of MT-ND3 transcription via FASTKD1 binding and CLPP’s processing of the mitoribosomal assembly factor MALSU1, which we establish as a disease gene. Through entropy analysis, we reveal unexpectedly heterogeneous protein-level variability across complexes and use complexome profiling to identify new complex-specific membership, including C15orf61’s association with complex V. We further observe substantial mtDNA copy number variation, notably upon disruption of the disease-related cobalamin biosynthesis protein MMADHC. Together, we establish new protein functions and provide a multilayered view into mitochondrial proteome regulation.

**Highlights:**

- Multiomic signatures across perturbations reveal extensive post-transcriptional regulation
- The TCA cycle enzyme MDH2 binds FASTKD1 to modulate MT-ND3 transcript levels
- MALSU1 is a CLPP protease substrate whose deficiency causes a mitochondrial disease
- C15orf61 binds ATP synthase and negatively regulates its higher order assembly
- MMADHC inversely affects mtDNA levels potentially mediated through LONP1

## INTRODUCTION

Mitochondria are evolutionary remnants of ancient proteobacteria that are essential for all eukaryotic life. Mitochondrial biogenesis involves a complex orchestration of protein translation, localization, import, processing, refolding, and organization of more than 1,000 proteins encoded by both the nuclear and mitochondrial genomes. Disruption to these processes, *e.g.* via genetic mutations or chemical perturbations, can cause proteostatic stress and cell death, demonstrating the need for precise and continuous calibration of mitochondrial protein content and stoichiometry.^1–4^ Moreover, given the long half-lives of many mitochondrial proteins,^5–7^ the need for rapid responses to changing cellular needs, and the subcellular specialization of these organelles,^8,9^ post-transcriptional means of regulation likely play a prominent role in mitochondrial maintenance and function.

Post-transcriptional regulation of protein abundance and activity comes in many forms. For example, cells exert precise control of protein complex subunit production to achieve proper stoichiometry;^10–12^ express various chaperones and proteases to assist protein folding, maturation, and turnover;^13,14^ and employ a vast array of protein-protein interactions to fine-tune enzymes and pathways.^15^ A prominent example within mitochondria are the oxidative phosphorylation (OxPhos) assembly factors — a diverse group of ∼70 proteins^16,17^ that assist the formation of complexes I-V and that are disrupted in a spectrum of primary mitochondrial diseases.^18,19^ Given that hundreds of mitochondrial protein functions are poorly defined,^20^ and that the molecular underpinnings of many mitochondrial diseases remain unclear,^21–23^ it is reasonable to expect that myriad regulatory protein activities remain to be discovered.

Approaches to define protein function and post-transcriptional regulation run the gamut from affinity-enrichment mass spectrometry^24–26^ to single-cell Perturb-seq^27^ to AI-driven computational predictions.^28^ However, systematic efforts that measure cellular responses to diverse perturbations across matched molecular classes (*e.g.*, transcripts, proteins, metabolites) hold key advantages.^29–31^ First, the multidimensionality of these approaches enables more refined, causal interpretations (*e.g.*, discerning that the elevation of a metabolite is due to an increase in the enzyme that produces it versus a decrease in the enzyme that consumes it).

Second, the breadth of contrasting perturbations (*e.g.*, gene knockouts) enables identification of unique gene-molecule relationships from among common or “noisy” cellular responses even when the effect is subtle (*e.g.*, discerning whether the metabolite increase above is unique to a specific gene disruption). Thus, this strategy, while labor-intensive, not only captures the perturbation-induced effect(s) but also catalyzes discovery of causal links between each perturbed gene and observed molecular fluctuations.

Here, we perform a deep multiomic analysis of more than 200 genomic perturbations to genes encoding mitochondrial proteins. This effort more than triples the molecular measurements of our original MITOMICS study^30^ by adding matched transcriptomic and mtDNA abundance analyses across all cell lines. These data, along with supporting affinity-enrichment and complexome profiling mass spectrometry experiments, enabled us to forge new connections between mitochondrial proteins and post-transcriptional processes. These findings include “moonlighting” activities for well-known metabolic proteins, new functions for uncharacterized proteins, and new substrates for a core protease. Moreover, we demonstrate the importance of these activities to human health by linking them to two mitochondrial diseases. Finally, we packaged our collection of >26 million molecular measurements of mitochondrial gene regulation and protein function into an interactive website (online upon peer-review) that will empower extensive basic and translational studies into the role of mitochondria in human health and disease.

## RESULTS

### Deep multiomic profiling of mitochondrial perturbations

Recently, we generated a deep multiomic dataset (MITOMICS) from a panel of 203 human HAP1 knockout (KO) cell lines, each with a single nucleus-encoded mitochondrial gene disruption.^30^ The targeted genes, representing more than 10% of the MitoCarta compendium,^17^ include representatives for most established mitochondrial pathways and many other encoding proteins that remain poorly characterized. The resulting molecular profiles have proven highly effective for suggesting new mitochondrial protein functions and linking their dysfunction to human diseases. Nonetheless, the MITOMICS dataset was limited to protein and metabolite measurements, thereby restricting key analyses, including those that seek to unveil discrete post-transcriptional protein functions.

Here, we substantially extend the MITOMICS dataset by adding matched transcriptomic profiles and mtDNA quantification data across all cell lines, and complexome profiling data on selected clones. These efforts more than tripled the number of molecular measurements and enable an extensive new range of analyses (**Fig. 1A**). In this study, we focus on three such analyses and the disparate discoveries they enable: 1) a nearest neighbor “outlier” analysis designed to identify molecular changes specific to a given KO, 2) an entropy analysis designed to distinguish dynamic pathways and complexes from those that remain static in response to genetic perturbations, and 3) an mtDNA copy number analysis that reveals the effect of each KO on mitochondrial genome abundance. Each analysis is bolstered by further biochemical and mass spectrometry (MS)-based experiments, including cross-linking affinity-enrichment MS (XL-AEMS) and complexome profiling. Collectively, these analyses create a new integrated MITOMICS dataset spanning genome dosage to transcript-protein relations to protein complex assembly.

**Figure 1.**
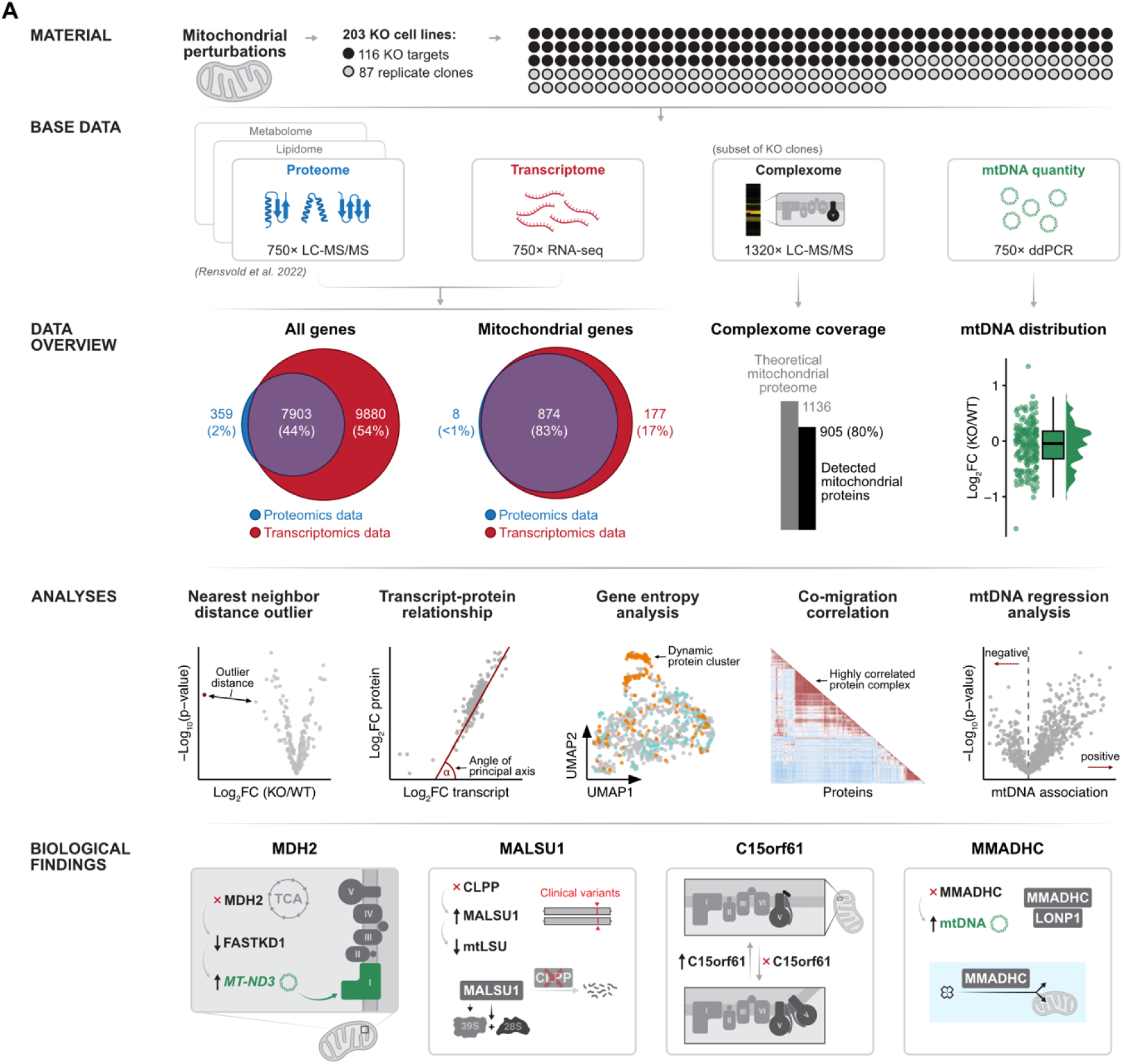
Study concept and data overview. (A) Study overview highlighting the obtained data types and performed analyses.

### Outlier analyses predict protein relationships

To begin expanding the MITOMICS dataset, we performed RNA-seq analyses on 3-4 independently cultured replicates from each cell line harvested in parallel with our original proteomics specimens. We attained high-quality measurements for 17,783 transcripts, including 7,903 genes (874 mitochondrial) with paired protein measurements across all 203 perturbations (**Fig. 1A**), making this, to our knowledge, the largest matched transcript-protein dataset across individual genetic perturbations.

From these extensive matched data, we made over 1.6 million protein-transcript comparisons, allowing us to estimate the extent to which gene expression alterations in our dataset were driven by transcriptional vs. post-transcriptional processes. Consistent with prior reports,^32–34^ we observe only a moderate overall protein-transcript correlation (**Fig. 2A**), though individual genes displayed substantial heterogeneity ranging from strongly positive to absent or slightly negative transcript-protein relationships (**Fig. S1A**). To quantify this uncoupling systematically, we calculated residuals from linear regression models and aggregated them by mitochondrial gene sets (**Fig. S1B, Table S1**). This analysis demonstrated that the abundances of protein complexes, particularly OxPhos components and the large mitoribosomal subunit (mtLSU), fluctuate predominantly at the protein level in response to our genetic perturbations, whereas intermediary metabolism pathways (*e.g.*, ketone metabolism) show tighter transcript-protein coupling following classical molecular dogma (**Fig. 2B**). The pronounced uncoupling was especially evident for mitochondrial genes (nuclear and mtDNA-encoded), which exhibited stronger proteomic but weaker transcriptomic variation compared to non-mitochondrial genes (**Fig. S1C**).

**Figure 2.**
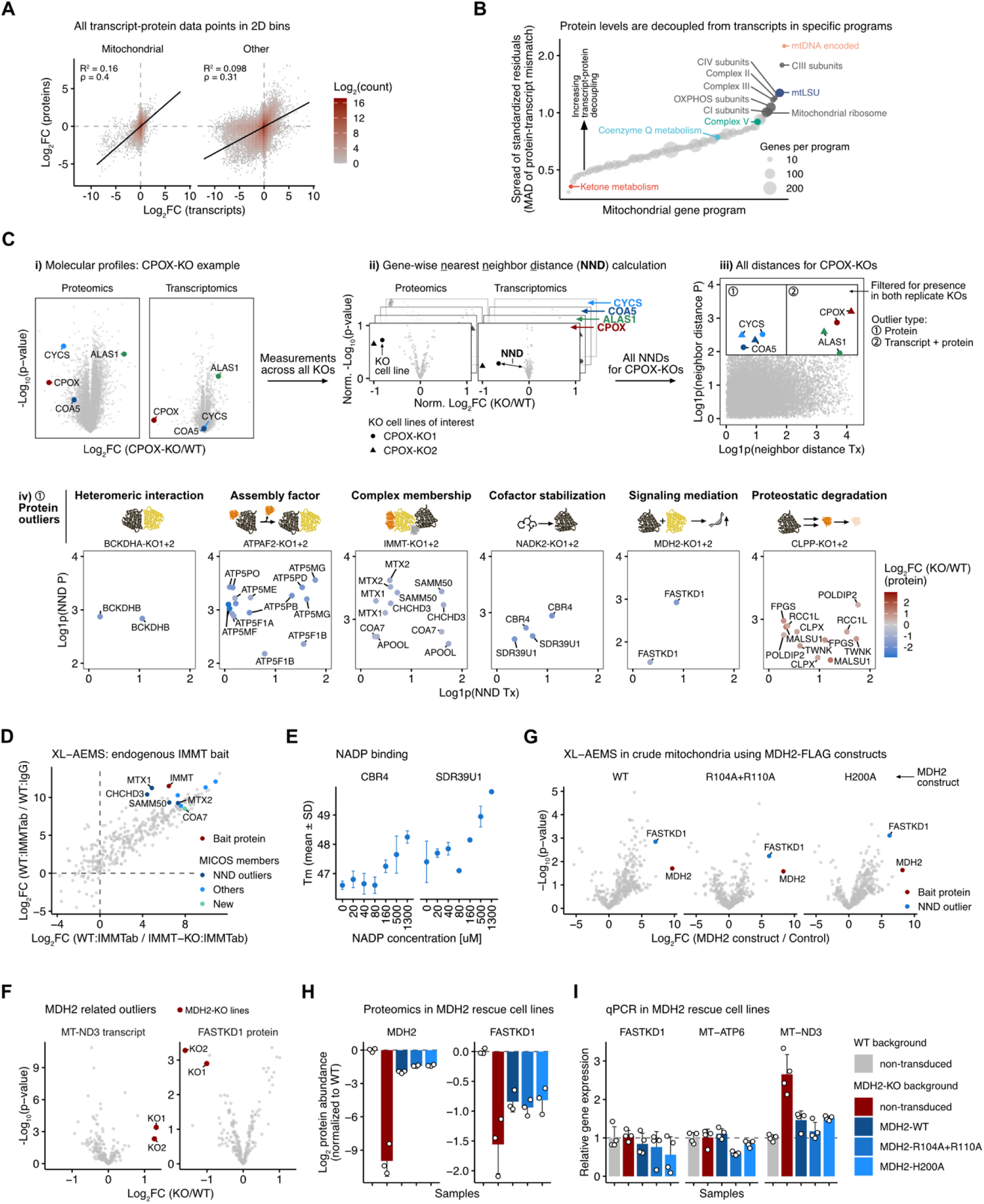
Nearest neighbor distance analysis predicts established and novel protein-protein relationships. (A) 2D histogram of log_2_ fold changes of transcript (x-axis) and protein (y-axis) levels across all knockout cell lines. The line and R-squared values are derived from a linear regression model. The correlation coefficients are based on Pearson correlation calculation. (B) Decoupling of transcript and protein changes. Linear regression residuals from transcript-protein relationships were calculated, grouped by mitochondrial gene sets and median absolute deviations were computed as a proxy for uncoupling of transcript-protein levels within a given mitochondrial gene set. (C) Nearest neighbor distance calculation concept, illustrated by the CPOX KO replicate clones. (i) Volcano plot of molecular profiles within one CPOX-KO cell line [cell line centric view; each dot is a molecule]. (ii) Volcano plots of specific molecules [gene centric view; each dot is a KO cell line]. Nearest neighbor distance is defined for each KO cell line across all molecules. (iii) All nearest neighbor distances for both CPOX KO cell lines, depicted as scatter plot representing transcript (x-axis) and protein (y-axis) distances. (iv) Example protein outliers of specific cell lines are depicted as individual plots. (D) Scatter plot of XL-AEMS experiment using an IMMT antibody to target the endogenous protein in wild-type mitochondria from HAP1 cells. (E) Melting temperatures of recombinant CBR4 and SDR39U1 proteins incubated with increasing concentrations of NADP. (F) Volcano plots of MT-ND3 transcript and FASTKD1 protein levels across all KO cell lines of the study. MDH2-KO cell lines are highlighted. (G) Volcano plots depicting XL-AEMS data using three different MDH2-FLAG constructs as bait compared to wild-type background. (H) MDH2-KO cell line was transduced with three different MDH2 constructs (wild-type, a catalytically inactive mutant (H200A), and a substrate-binding mutant (R104A+R110A)) and proteomics was performed. Bar plots of MDH2 and FASTKD1 are shown. Error bars represent standard deviation. (I) Quantitative qPCR in the same MDH2 stable cell lines targeting three different transcripts. Error bars represent standard deviation.

These results suggest heavy reliance on post-transcriptional regulation for most mitochondrial proteins — a phenomenon that can be explored via a nearest neighbor “outlier” analysis. To do so, we first plotted the effect that each targeted gene disruption (*e.g.*, *CPOX,* coproporphyrinogen oxidase; involved in heme biosynthesis) has on all measured proteins and transcripts (**Fig. 2Ci**). Next, we sequentially compared all such protein and transcript effects across all gene KOs and calculated nearest-neighbor distances (**Fig. 2Cii, Table S2**). These distances highlight outlier KO gene-protein and KO gene-transcript relationships — those changes that are particularly strong in one KO line (here the CPOX KO) versus all others, thus suggesting biological relationships between the KO gene and interrogated transcripts and proteins. Finally, by plotting CPOX KO-mediated protein-specific outliers vs protein+transcript outliers we can distinguish transcriptional (upper right quadrant) from post-transcriptional (upper left quadrant) relationships (**Fig. 2Ciii**).

As expected, CPOX KO lines displayed pronounced depletion of CPOX gene expression at both transcript and protein levels (upper right quadrant). Conversely, *ALAS1* (5-aminolevulinate synthase 1) — the upstream rate-limiting enzyme of heme biosynthesis — showed marked upregulation in CPOX KO cells at both the protein and transcript levels, likely due to relief of heme-mediated feedback inhibition.^35,36^ Notably, this same perturbation produced protein-specific outliers for CYCS (cytochrome c) and COA5 (cytochrome c oxidase assembly factor 5), consistent with their destabilization following heme *c* and heme *a* depletion, respectively. These results thus recapitulate established relationships between CPOX and other heme-related proteins and mirror observations in porphyria patients^37^ and ferrochelatase inhibition experiments.^38^ Moreover, these results demonstrate that this analysis can reveal multiple distinct protein-transcript and protein-protein relationship types.

We extended this analysis dataset-wide focusing on the top-left quadrant of the nearest-neighbor distance plot for each gene KO (**Fig. 2Civ**). This cleanly recovered various established protein-protein relationships, such as the heteromeric interaction between BCKDHA-BCKDHB of the branched-chain α-ketoacid dehydrogenase complex; ATPAF2’s role in maintaining and assembling ATP synthase subunits (**Fig. S2A**);^39^ and IMMT’s interaction with other members of the MICOS complex (**Fig. 2D**).^40,41^ Additionally, multiple unexpected relationships became evident. In the latter example, COA7 (cytochrome *c* oxidase assembly factor 7) emerged as a strong outlier with IMMT, which we validated by endogenous co-immunoprecipitation (**Fig. S2B**), potentially indicating an integration of OxPhos assembly with MICOS function. We further discovered that loss of NADK2, which generates NADP(H) from NAD(H), specifically destabilized CBR4 and SDR39U1. These two short-chain dehydrogenases^42^ are reported to bind NADP(H) *in vitro* (PDB: 4CQM, 4B4O and ref^43^), which we confirmed by differential scanning fluorimetry using purified enzymes (**Fig. 2E**). Determining endogenous NAD(H) vs. NADP(H) cofactor selectivity is a well-documented difficulty.^44^ Here, CBR4 and SDR39U1 are the only two proteins among the 22 mitochondrial proteins in our dataset annotated to bind NADP(H)^45^ that showed exceptional NADK2-dependent stability (**Fig. S2C**), suggesting a specialized dependency for NADK2-generated NADP(H). These data provide a valuable clue to SDR39U1’s unknown function and, for CBR4, complement recent findings linking NADK2 to mitochondrial fatty acid synthesis.^46^

Our outlier analyses revealed a particularly intriguing new relationship for the TCA cycle enzyme MDH2 (malate dehydrogenase 2). Among all KO cell lines, MDH2 KOs clearly had the highest abundance of the MT-ND3 transcript (whose protein was undetected) (**Fig. 2F, S2D**).

Moreover, these same MDH2 KO lines were outliers for depletion of the RNA-binding protein FASTKD1 (**Fig. 2F**) — a protein linked to suppression of MT-ND3 expression.^47^ To explore this further, we performed cross-linking affinity-enrichment mass spectrometry (XL-AEMS) in WT and MDH2 KO lines targeting the endogenous MDH2 protein, and identified a robust interaction of FASTKD1 with MDH2 (**Fig. S2E,F**). We generated an alternative system re-expressing wild-type or catalytic mutant MDH2 constructs in the MDH2 KO background (**Fig. S2G**), demonstrating that this interaction is stronger than known MDH2 interactors^48,49^ and that it persists independently of MDH2’s enzymatic activity (**Fig. 2G, S2H**). Moreover, the same rescue cell lines showed partial correction of FASTKD1 protein loss and near-complete normalization of MT-ND3 transcript levels (**Fig. 2H,I, S2I**). These results demonstrate that MDH2 exerts a marked effect on MT-ND3 levels in a manner distinct from its malate dehydrogenase activity, suggesting that MDH2 may help coordinate TCA cycle activity with OxPhos regulation via mitochondrial transcript stability. More broadly, this finding bolsters the growing notion that well-established proteins can possess auxiliary “moonlighting” functions based on cellular context.^50^

Beyond these findings, our outlier analyses suggest many more protein-protein relationships yet to be explored. These include suggested protease substrates, links between mitochondrial type II fatty acid synthesis and Fe-S cluster generation, connections between coenzyme Q production and mitochondrial protein import, and various unannotated relationships between disparate individual proteins (**Fig. S3A**). Together, these examples demonstrate that our approach effectively recovers established biology and reveals novel relationships.

### MALSU1 dysregulation causes disease

CLPP KO cells exhibited the most extensive array of protein outliers (**Fig. 2A**), consistent with CLPP’s role as a mitochondrial matrix protease linked to protein quality control and homeostasis,^51^ and to Perrault syndrome when mutated. These outliers include multiple established CLPP substrates and a selection of unexpected hits. To test these interactions, we again performed XL-AEMS using the substrate-trapping S153A mutant of CLPP (**Fig. 3A**). We recovered several of CLPP’s nearest neighbor outliers, including CLPX (the cognate AAA^+^ unfoldase),^52^ POLIDP2 (a heme-sensing CLPX adaptor),^53^ and the newly proposed substrates FPGS and MALSU1 — a poorly characterized mitoribosome assembly factor (**Fig. 3B**).

**Figure 3.**
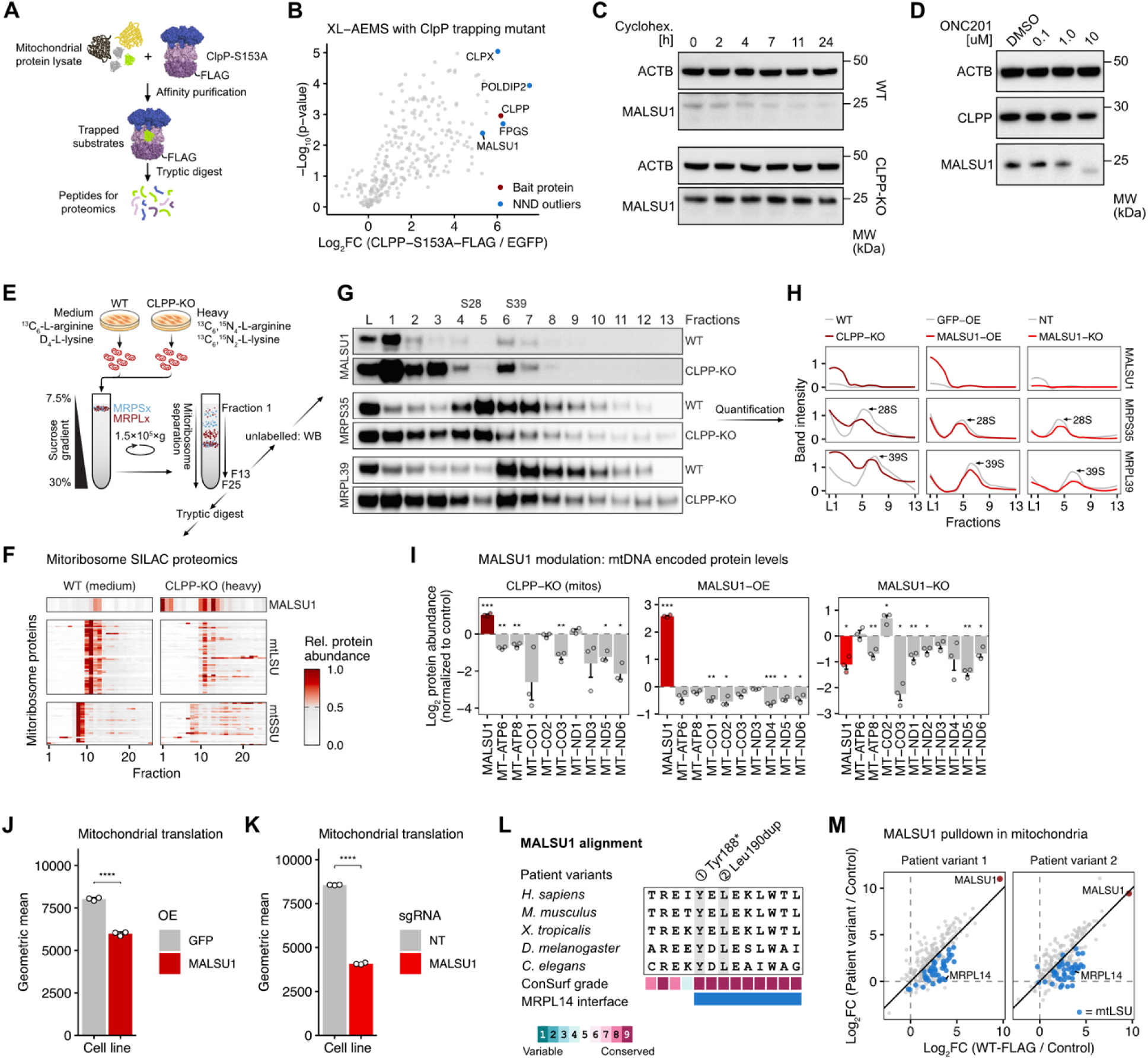
MALSU1 is a CLPP substrate and causes a mitochondrial disease. (A) Scheme of trapping pulldown method using the S153A trapping mutant of CLPP. (B) Volcano plot based on XL-AEMS experiment using the CLPP-S153A mutant. Highlighted proteins are the CLPP bait protein and the predicted outliers from the nearest neighbor distance analysis. (C) Western blot for MALSU1 based on HAP1 cells treated with cycloheximide for 24 hours. (D) Blot for CLPP and MALSU1 following treatment with the CLPP activator ONC201 at increasing concentrations for 24 hours. (E) Scheme of mitoribosome sedimentation ultracentrifugation assay. (F) Heatmap of mitoribosome proteins across all fractions from a SILAC-based mitoribosome sedimentation experiment with proteomics readout, using HAP1 wild-type and CLPP-KO cells. (G) Western blot based on 13 fractions from a mitoribosome sedimentation expeirment, blotting for exemplary mtLSU and mtSSU proteins as well as MALSU1. (H) Western blot band quantification from mitoribosome sedimentation experiments using CLPP knockout, MALSU1 overexpression or knockout cells. (I) Data from three different proteomics experiments using samples from crude mitochondria extracted from HAP1 CLPP-KO cells, cell lysates from MALSU1 overexpressing, or knockout cells (from left to right). Statistical significance was assessed using Welch’s t-test; p-values ≤0.05 (*), ≤0.01 (**), ≤0.001(***). (J) Mitochondrial translation assay based on L-homopropargylglycine incorporation, performed in 293T cells overexpressing GFP or MALSU1. Statistical significance was assessed using Welch’s t-test; p-values ≤0.0001(****). (K) Mitochondrial translation assay performed in 293T CRISPR/Cas9-MALSU1-KO cells, compared to cells transduced with a non-targeting guide. Statistical significance was assessed using Welch’s t-test; p-values ≤0.0001(****). (L) MALSU1 amino acid sequence alignment based on selected species using the Clustal Omega MSA tool.^113^ ConSurf scores were calculated based on PDB: 7OF0.^114^ (M) XL-AEMS scatter plot comparing MALSU1 wild-type and patient variant interactomes. Mitoribosome proteins are highlighted in blue, MALSU1 in red. The bold line represents the diagonal where proteins with equal enrichment in both samples (wild-type and patient variant) would fall.

We consistently observed cellular MALSU1 accumulation when we depleted CLPP across multiple systems, including HAP1 knockouts, CRISPRi-mediated depletion in 293T and U2OS cells (**Fig. S4A**), and siRNA knockdown in 293T cells (**Fig. S4B**). In rescue experiments using CLPP KO cells, we found that only wild-type CLPP, but not a catalytically inactive S153A mutant, restored normal MALSU1 levels (**Fig. S4C**). Through cycloheximide chase experiments, we observed near-complete MALSU1 degradation in wild-type cells after 24 hours, while MALSU1 remained stable in CLPP KO cells (**Fig. 3C**). Conversely, when we enhanced CLPP activity either genetically (via the constitutively active Y118A mutant,^54^ **Fig. S4D**) or pharmacologically (using the imipridone compound ONC201,^55^ **Fig. 3D**), we detected reduced MALSU1 levels, establishing MALSU1 as a bona fide CLPP substrate.

Recently, MALSU1 has been linked to mitoribosome assembly by promoting large subunit (mtLSU) maturation and integration with the small subunit (mtSSU).^56–58^ Our connection of MALSU1 to CLPP suggests that careful calibration of MALSU1 levels might be important for mitoribosome function, and that MALSU1 could act as a key factor in Perrault syndrome pathogenesis. In support of this, CLPP-deficient cells displayed reduced oxygen consumption (**Fig. S4E**) and diminished mtDNA-encoded protein levels in both siRNA-treated 293T cells (**Fig. S4F**) and HAP1 CLPP KO lines (**Fig. S4G**), although these phenotypes could also be influenced by other CLPP substrates. Notably, these phenotypes were present despite sufficient mtDNA transcript levels and modest increases in mitoribosome proteins (**Fig. S4G,H**), leading us to more closely study the assembly of mitoribosomes using a sedimentation assay (**Fig. 3E**). We identified decreased 28S and 39S subunit formation in CLPP KO cells by proteomics and immunoblotting (**Fig. 3F,G**). Critically, direct MALSU1 modulation, either overexpression or knockout, recapitulated this assembly defect (**Fig. 3H**), accompanied by reduced mtDNA-encoded proteins (**Fig. 3I**) due to impaired mitochondrial translation as measured by L-homopropargylglycine incorporation (**Fig. 3J,K**). Similar examples of such a “Goldilocks” phenomenon, in which either too much or too little protein expression is detrimental, has been reported for ribosomal factors like BOP1^59^ or coenzyme Q (CoQ) proteins such as Coq5 in yeast.^60^

The clinical relevance of MALSU1 regulation emerged through our discovery of a patient harboring bi-allelic variants (p.(Tyr188*);p.(Leu190dup)) presenting with exercise intolerance, short stature, and elevated lactate (identified through GeneMatcher,^61^ personal communication R. Rodenburg). Both variants mapped to highly conserved residues/regions (**Fig. 3L**) and, when expressed in MALSU1 KO cells, failed to maintain interaction with the direct binding partner MRPL14 (PDB: 7OF0) and other mtLSU components (**Fig. 3M**), supporting their pathogenicity. These findings establish MALSU1 dysregulation downstream of CLPP deficiency as a mechanism underlying mitoribosome dysfunction and the likely driver of a previously unrecognized mitochondrial disease. More broadly, these results further demonstrate the power of our outlier analyses to reveal key mitochondrial protein-protein relationships important for human health.

### Transcript-protein relationships differ between pathways

The analyses above focus primarily on proteins whose levels are not strongly predicted by their corresponding transcript abundance. However, many other proteins are well correlated with their transcripts. To assess these differing relationships on a gene-by-gene basis, we performed a principal component analysis of the transcript-protein space for each gene. This method calculates the angle of the first principal component (PC1) as a representative metric of either transcription-driven regulation (45-degree angle) (**Fig. 4A, S5A, Table S3**) or protein-level regulation (90-degree angle), with the variance explained by PC1 quantifying the relationship strength. We applied this approach to all 7,903 shared genes to predict general regulatory modes for defined gene sets.

**Figure 4.**
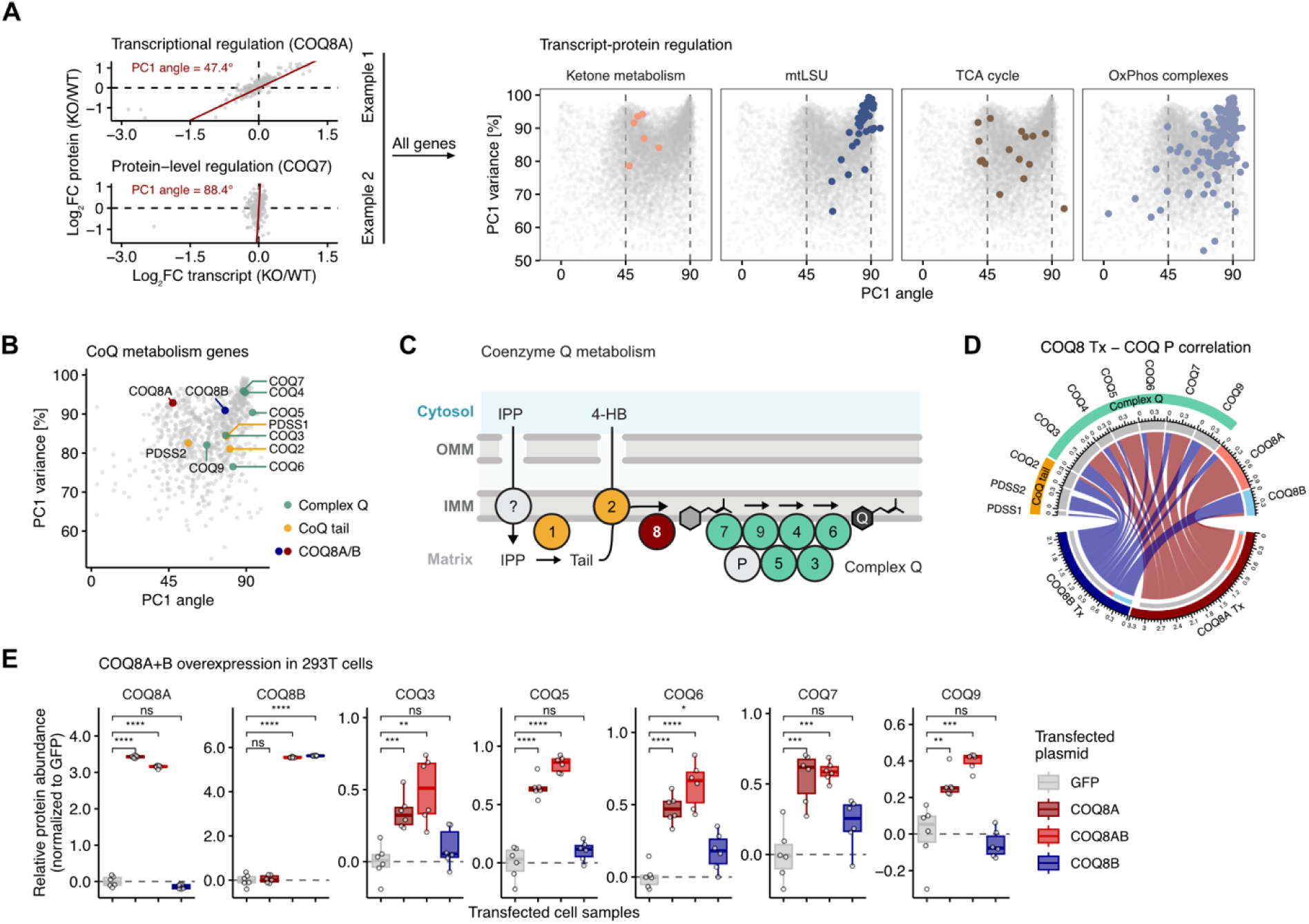
Gene-wise molecular regulation defined by principal component analysis. (A) Left: Scatter plot indicating the transcript and protein log^2^ fold changes of the two example genes. The red line represents the first principal component (PC1). Right: PC1 metrics (angle, variance explained) are plotted for all genes in the dataset with shared transcript-protein information; selected mitochondrial gene sets are highlighted. (B) Scatter plot of PC1 metrics filtered for mitochondrial genes. Highlighted genes belong to the coenzyme Q_10_ (CoQ) biosynthesis pathway. (C) Schematic drawing of the CoQ biosynthetic pathway. The numerals refer to the individual *COQ* genes. (D) Chord plot depicting the correlative relationships of COQ8A and COQ8B transcripts to all detected protein members of the CoQ gene set. Connector thickness corresponds to Spearman correlation coefficient. (E) Box plots showing proteomics results for CoQ proteins, following overexpression of COQ8A and/or COQ8B. Statistical significance was assessed using Welch’s t-test; p-values ≤0.05 (*), ≤0.01 (**), ≤0.001(***), ≤0.0001(****).

Within mitochondrial pathways, transcripts and proteins related to intermediary metabolism show strong correlation, while protein complexes display more prominent protein-level regulation patterns (**Fig. 4A**), consistent with our transcript-protein uncoupling metric (**Fig. 2B**). Interestingly, some pathways show strong overall protein-level regulation but feature one or few members with high transcript-level correlation. For example, within the coenzyme Q (CoQ) biosynthesis pathway, PDSS2 shows high transcript correlation, but its binding partner PDSS1 exhibits more protein-level regulation (**Fig. 4B**). This disparate control is interesting given that PDSS1/2 form an obligate heterotetramer to synthesize the CoQ tail,^62^ and thus their ultimate stoichiometry needs to be 1:1. Notably, mutations in either gene are sufficient to cause primary CoQ deficiency.^63,64^ Similarly, COQ8A, but not its paralog COQ8B, exhibits strong transcript-level regulation compared to other proteins that form the complex Q metabolon (**Fig. 4B**).

COQ8A/B function as atypical kinases/ATPases,^65^ and partially rescue induced complex Q proteins loss in yeast.^66^ Since COQ8A and COQ8B mutations cause primary CoQ deficiency associated with severe neurological or kidney disorders in humans, respectively,^67,68^ understanding their role in the hierarchical regulation of the CoQ pathway warrants investigation (**Fig. 4C**). To that end, we next performed transcript-protein correlation analyses that suggested COQ8A may play a higher-level role in regulating the CoQ metabolon within these cells (**Fig. 4D, Fig. S5B,C**). COQ8A overexpression increased CoQ metabolon proteins independently of COQ8B, supporting these predictions (**Fig. 4E**). Collectively, these data support the notion that disparate regulatory processes exist between and within mitochondrial pathways to titrate protein abundance, potentially with clinical relevance (**Table S3**).

### C15orf61 negatively regulates ATP synthase

Our analyses thus far were effective at identifying diverse types of protein relationships, but they depended on proteins of interest exhibiting dynamic profiles to forge these connections. We next assessed which proteins exhibited such dynamic profiles and which instead were largely static in response to our genetic perturbations, reasoning that the latter may possess distinct forms of regulation. To quantify this variability, we calculated an entropy metric for each gene (similar concepts used in refs^69,70^; more details in methods). Focusing on the protein data, we mapped entropy scores onto mitochondrial protein embeddings, which simultaneously clustered proteins with similar expression patterns and revealed their characteristic (static vs. dynamic) variability signatures (**Fig. 5A, Table S3**). As an example, the large mitoribosome subunit (mtLSU) exhibited high dynamics, consistent with highly responsive protein-level regulation mediated by (among other mechanisms) CLPP proteostasis as shown above (**Fig. 2C, 3**).

**Figure 5.**
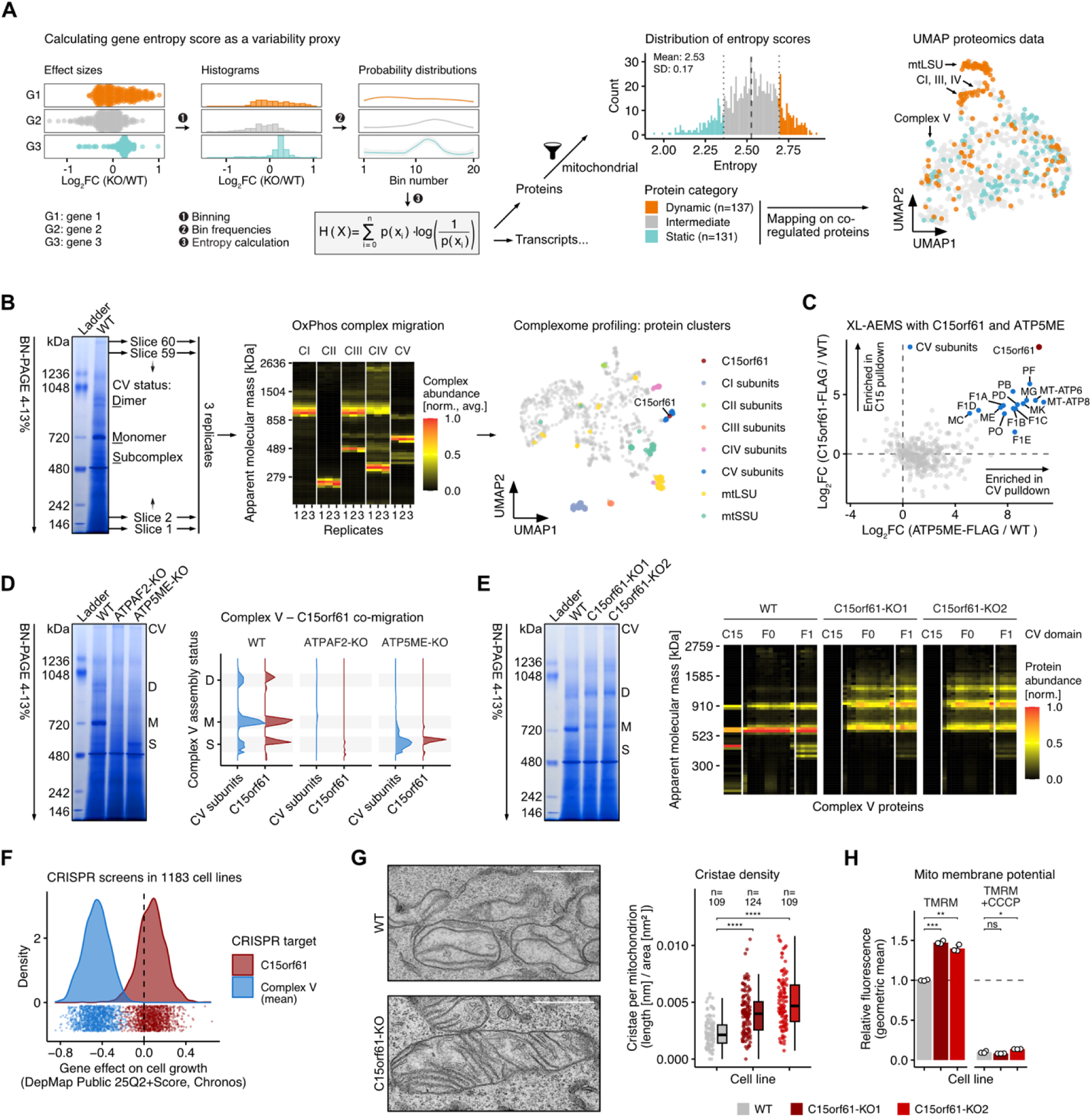
Identification of C15orf61 as complex V subunit and cellular and organellar consequences of its loss. (A) Entropy-based variability classification of genes across transcriptomics and proteomics, which is mapped onto a UMAP-based molecule embedding (protein data shown). (B) Complexome profiling workflow using blue native polyacrylamide gel electrophoresis (BN-PAGE) and subsequent proteomics across 60 slices per lane, performed in triplicates. Data analysis by heatmap depicting normalized average abundances for OxPhos complexes or UMAP to identify proteins with similar migration patterns. (C) Dual XL-AEMS experiment using two different bait proteins (C15orf61-, and ATP5ME-FLAG). Known CV subunits and C15orf61 are highlighted. (D) Complexome profiling of mitochondria extracted from HAP1 wild-type, ATPAF2-, and ATP5ME-KO cells. The density plots represent the average migration of CV proteins, and C15orf61. (E) Complexome profiling of mitochondria extracted from HAP1 wild-type, and two replicates of C15orf61-KO cells. Heatmaps depict individual CV proteins in columns. (F) Density plot depicting DepMap growth scores for CRISPR knockouts in CV genes (averaged) compared to C15orf61. (G) Representative transmission electron microscopy images from wild-type and C15orf61-KO cells. Individual mitochondria were quantified in each cell line to assess cristae density, as represented by single dots and box plot summaries. Statistical significance was assessed using Welch’s t-test; p-values ≤0.0001(****). (H) Bar plot depicting mitochondrial membrane potential measured by tetramethylrhodamine methyl ester (TMRM) fluorescence (geometric mean from fluorescence-activated cell sorting experiment). Carbonyl cyanide 3-chlorophenylhydrazone (CCCP) was used as protonophore control. Statistical significance was assessed using Welch’s t-test; p-values ≤0.05 (*), ≤0.01 (**), ≤0.001(***).

Conversely, ATP synthase (complex V, CV) subunits remained remarkably static, suggesting CV adaptation, if present in our cell lines, occurs through mechanisms beyond protein abundance changes (**Fig. 5A**).

We hypothesized that complexome profiling — a gentle and comprehensive method for identifying protein interactions^71,72^ — could identify modulators that affect overall CV architecture without altering CV subunit levels. Exploring CV modulators seemed particularly relevant given the limited mechanistic understanding of ATP synthase multimerization.^73,74^ To begin, we carefully optimized mitochondrial lysis and subsequent BN-PAGE separation for detection of CV assembly states. Our subsequent profiling resolved ∼900 mitochondrial proteins and captured all CV assembly states along with most other OxPhos and mitochondrial complexes (**Fig. 5B, Table S4**). Using UMAP-based analysis, we identified the uncharacterized protein C15orf61 as a non-canonical CV component, showing the highest migration correlation among non-CV proteins with CV subunits (**Fig. 5B, S6A**) — a finding that we corroborated across three independent complexome datasets (**Fig. S6B**).^75,76^

We next performed separate XL-AEMS experiments on FLAG-tagged constructs of C15orf61 and the CV subunit ATP5ME, which confirmed this physical interaction (**Fig. 5C**). Crosslinked peptides mapped the interaction of C15orf61 to the CV c-ring (encoded by *ATP5MC1-3*) (**Fig. S6C**), which was further supported through a nearly congruent co-migration alignment between C15orf61 and ATP5MC (**Fig. S6D,E**), near-complete C15orf61 loss in *ATP5MC1-3* triple knockout cells (**Fig. S6F**), and the inner mitochondrial membrane localization of C15orf61 determined by a proteinase K assay (**Fig. S6G**). We further profiled complexomes of ATPAF2-KO and ATP5ME-KO cells, revealing C15orf61 association with all CV assembly states (dimers, monomers, and subcomplexes) (**Fig. 5D, Table S4**), suggesting that C15orf61 does not operate as a classical assembly factor.

While C15orf61 knockout had no consistent effect on CV subunit transcript or protein abundances (**Fig. S6H**), we observed substantial redistribution of CV toward dimers and oligomers at the expense of monomers in C15orf61-KO mitochondria (**Fig. 5E, Table S5**). We detected the opposite effect upon C15orf61 overexpression in HAP1 cells (**Fig. S6I, Table S6**). While CV multimerization changes did not alter CV’s ATPase activity (**Fig. S6J**), C15orf61 loss increased cell proliferation across >1,000 DepMap experiments (**Fig. 5F**), increased cristae density (**Fig. 5G**), enhanced mitochondrial network connectivity (**Fig. S6K**), and elevated mitochondrial membrane potential (**Fig. 5H**). Together, these findings establish C15orf61 as a supernumerary CV subunit that negatively regulates multimerization, and we propose renaming it ATP5MM (ATP synthase membrane-associated multimerization modulator).

### MMADHC links cobalamin metabolism to mtDNA maintenance

Mitochondrial proteins are encoded by two genomes, adding an additional layer of regulatory complexity absent for other cellular compartments. A third means of modulating mitochondrial protein architecture is thus to modify mitochondrial genome maintenance. To explore potential connections between our targeted genes and mitochondrial genome dosage, we used digital droplet PCR (ddPCR) to measure mtDNA content across all knockout lines. By quantifying mitochondrial (MT-ND1) and nuclear (B2M) gene abundances in purified DNA samples (**Fig. 6A, Table S7**), we found 20% of lines (N=43) exhibited reduced mtDNA, while 7% (N=15) showed increased mtDNA content (**Fig. 6A**). From this data, we first tested whether mtDNA abundance predicts mitochondrial transcript and protein levels using linear regression (**Table S7**). Focusing on mtDNA-encoded genes, we found that transcripts showed no association, while a subset of the six detected mtDNA-encoded proteins correlated with genome copy number (**Fig. 6A, S7A**), reinforcing our previous observation of a predominant protein-centric regulation mode for these polycistronically transcribed genes (**Fig. 2B**).

**Figure 6.**
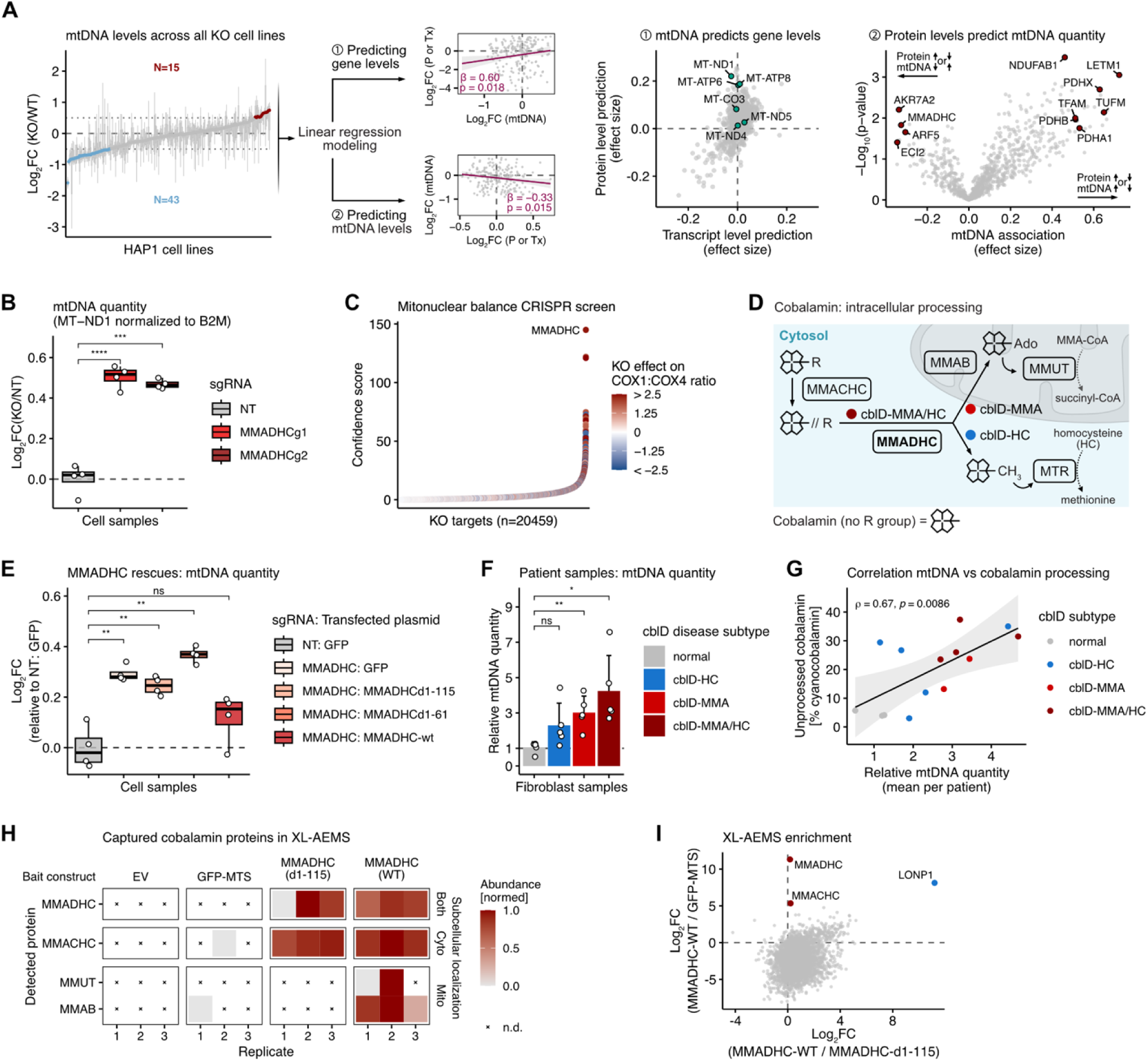
mtDNA association studies reveal inverse relationship of MMADHC and mtDNA quantity. (A) Levels of mtDNA quantity were determined using ddPCR across all HAP1 KO cell lines and compared to WT values. Linear regressions were performed in both directions (mtDNA ∼ molecule and molecule ∼ mtDNA) to identify associations between mtDNA quantity and transcript/protein levels. (B) qPCR-based mtDNA quantification in 293T CRISPR/Cas9-KO cells. NT, non-targeting. (C) Ranked knockout genes from a mito-nuclear CRISPR screen based on confidence scores. Color gradient indicates the ratio of a mtDNA-encoded to a nuclear-encoded complex IV subunit. (D) Scheme of intracellular cobalamin processing focusing on the cytosolic-mitochondrial branch point involving the MMADHC protein. (E) qPCR-based mtDNA quantification in MMADHC-KO cells rescued with different MMADHC constructs. Statistical significance was assessed using Welch’s t-test; p-values ≤0.01 (**). (F) mtDNA levels in cblD patient fibroblasts determined by qPCR. Points represent mean values for one patient cell line, assessed in four technical replicates. Statistical significance was assessed using Welch’s t-test; p-values ≤0.05 (*), ≤0.01 (**). (G) Correlation between mtDNA abundance from and biochemical cobalamin pathway activity in patient fibroblasts (subset with available data). (H) XL-AEMS experiment using two different MMADHC constructs and an empty vector (EV) and GFP-MTS control. Selected proteins are shown; abundance values were z-transformed. MTS, mitochondrial targeting sequence. (I) XL-AEMS comparing wild-type MMADHC interactors against cytosolic MMADHC (d1-115) and non-specific mitochondrial binding (MTS-GFP control).

We then performed the reverse calculation to identify proteins whose abundances predict mtDNA levels (**Table S7**). As expected, TFAM emerged as one of the strongest positive predictors, validating its role in mtDNA stabilization,^77^ alongside unexpected proteins including subunits of the pyruvate dehydrogenase complex. Conversely, we identified negative associations predicting mtDNA elevation upon protein depletion. We validated several candidates (**Fig. S7B,C**) and prioritized MMADHC, which showed robust mtDNA increase upon knockout in two independent clones (**Fig. 6B, S7D**). Interestingly, analysis of external datasets corroborated this relationship: re-examining data from a CRISPR screen on mito-nuclear balance^78^ revealed elevated mtDNA-encoded COX1 (MT-CO1) relative to nuclear-encoded COX4 following MMADHC knockout (**Fig. 6C**), and analyzing data from a genome-wide single-cell CRISPRi screen^27^ uncovered increased mtDNA transcripts upon MMADHC knockdown (**Fig. S7E**) — relationships not reported in either original study.

MMADHC mediates intracellular cobalamin trafficking at the cytosolic-mitochondrial branch point, directing cobalamin to either cytosolic methionine synthase (MTR) or mitochondrial methylmalonyl-CoA mutase (MMUT) (**Fig. 6D**). MMADHC deficiency causes three clinically and biochemically distinct diseases characterized by homocysteine accumulation, methylmalonic acidemia, or both, depending on the variants’ impact on the mitochondrial targeting sequence.^79–81^ We hypothesized the mtDNA phenotype requires mitochondrial MMADHC function and confirmed this through rescue experiments: full-length MMADHC rectified mtDNA levels, whereas constructs lacking the mitochondrial targeting sequence failed to do so (**Fig. 6E**). We examined fibroblasts from patients representing all three disease phenotypes (**Fig. 6D**) and found that only MMADHC defects affecting its mitochondrial protein function exhibited elevated mtDNA (**Fig. 6F, Fig. S7F**). Furthermore, we observed strong correlation between impaired cellular cobalamin processing and mtDNA abundance in these patient cell lines (**Fig. 6G**).

Finally, we identified mitochondria-specific MMADHC interactors by controlling for non-specific mitochondrial binding (MTS-GFP) and subtracting cytosolic interactions (MMADHC-d1-115) (**Fig. 6H**), revealing LONP1 as the singularly prominent interaction partner (**Fig. 6I**). This result is intriguing given that LONP1 controls mtDNA homeostasis through proteostatic regulation of TFAM.^82^ These findings unexpectedly establish MMADHC as a regulator of mtDNA maintenance, likely mediated by its direct interaction with LONP1.

### Integrative MITOMICS analyses

The various analyses above demonstrate how our MITOMICS data can help bolster and extend previous findings and reveal new relationships between mitochondrial genes and proteins.

When paired with orthogonal methodologies — *e.g.*, the complexome profiling, XL-AEMS proteomics, or mtDNA quantification we leverage here — these approaches can predict and validate mitochondrial protein functions, reveal how individual factors shape mitochondrial proteome architecture, and connect gene disruptions to human disease.

Beyond what we show here, our data will enable extensive future investigations, including those focusing on non-mitochondrial genes, genes with only transcript data, or non-coding RNAs. Our online resource integrates all data collected from our MITOMICS studies, features ready-made interfaces for the analyses described in this work, and offers all our >26 million molecular measurements in downloadable files. Our data, especially when combined with other substantial multiomic resources, promises to accelerate basic mitochondrial investigations and efforts to link mitochondrial dysfunction to human disease.

## DISCUSSION

Human mitochondria contain over a thousand proteins whose levels must be precisely balanced to enable their functional cooperation. Despite this, systematic efforts to elucidate mechanisms governing orchestrated proteome expression and function have lagged behind those aimed primarily at cataloging proteome membership.^83^ Several factors complicate the former investigations: approximately one-fifth of proteins with confirmed mitochondrial localization remain functionally uncharacterized,^20^ many mitochondrial proteins exhibit moonlighting behavior,^84–86^ and deciphering these detailed orchestration mechanisms requires sophisticated experimental and analytical systems. To address these challenges, we developed a multimodal approach integrating mtDNA quantification with transcriptomic, proteomic, and complexome profiling across multiple genetic perturbations to systematically analyze mitochondrial proteome regulation and function.

Our work reveals a heavy reliance of mitochondrial genes on protein-centric regulatory mechanisms, though some pathways employ both transcript- and protein-driven processes, suggesting hierarchical regulatory organization. We further probed transcript and protein activities for each gene via our nearest neighbor outlier analyses to establish concrete relationships, recapitulating known biology and uncovering new findings centered on two themes: moonlighting functions and precise protein-level control. We identified MDH2 as a new molecular link between the TCA cycle and oxidative phosphorylation. Beyond its established enzymatic role in the TCA cycle, MDH2 appears to function structurally by modulating FASTKD1, thereby potentially influencing complex I levels directly. This finding is reminiscent of aconitase’s auxiliary function in yeast mtDNA maintenance^87^ and of the growing list of metabolic enzymes found to bind RNA.^88^ Together with our findings here, these examples suggest that extensive integration of metabolic signals with gene expression may be driven by such moonlighting functions. We also identified multiple CLPP substrates, most notably MALSU1, whose pronounced accumulation in CLPP-deficient cells likely establishes it as a key pathomechanistic factor underlying Perrault syndrome. Moreover, our discovery that MALSU1 variants likely cause mitochondrial disease independently underscores the critical importance of precisely controlling its protein levels. Of note, we did not detect ERAL1 as a CLPP substrate despite previous reports linking this protein to mitoribosome defects in CLPP deficiency,^89^ suggesting that CLPP-mediated protein regulation may exhibit cell-type or tissue-specific dependencies. We propose additional candidate substrates that likely require similar precision in protein-level control for mitochondrial health.

An unexpected benefit of our approach was its ability to highlight proteins displaying minimal variability across perturbations, suggesting regulatory mechanisms independent of protein-level control. Indeed, ATP synthase, which was among the most static complexes in our study, exhibited the clearest new interacting partner through complexome profiling: C15orf61 (ATP5MM), a finding that is corroborated by other complexome analyses from distinct cell types.^90^ Rather than affecting ATP synthase protein levels, ATP5MM appears to modulate complex multimerization. ATP synthase dimers are known to localize to cristae tips^91,92^ — invaginations of the inner mitochondrial membrane (IMM) — where they both generate ATP leveraging the proton-motive force and contribute structurally to cristae formation and maintenance. Recent cryo-electron tomography has revealed detailed resolution of in-situ dimer rows along cristae tips,^93^ and it is thought that dimerization can drive IMM remodeling.^94,95^ The discovery of ATP5MM suggests that cells possess proteins that regulate mitochondrial shape and metabolic state through modulation of ATP synthase multimerization. While our results localize ATP5MM to the IMM and demonstrate clear molecular and cellular consequences of its loss, future work must establish its precise binding interface with ATP synthase, interaction stoichiometry and occupancy, and the protein-level mechanisms underlying its functional effects.

Protein homeostasis in mitochondria can also be achieved through modulation of mtDNA abundance. Prior work has found associations between nuclear loci and mtDNA copy number^96^ and demonstrated the importance of coordinating proteins that originate from either the nuclear or mitochondrial genome but participate in the formation of the same OxPhos complex.^78,97^ This coordination mirrors the mechanism employed by FASTKD5, which regulates mt-mRNA stability,^98^ analogous to our discovery of FASTKD1’s dependence on MDH2. By integrating mtDNA abundance measurements with our multimodal dataset, we unexpectedly identified MMADHC as a negative regulator of mtDNA levels. MMADHC serves as a chaperone in the intracellular cobalamin (B12) pathway, facilitating cytosolic B12 delivery to methionine synthase and mitochondrial B12 delivery to methylmalonyl-CoA mutase. While the mechanics of this delivery remains elusive,^99^ mitochondrial MMADHC dysfunction causes an inborn error of metabolism characterized by methylmalonic acid accumulation. Interestingly, our data reveal that mtDNA levels concomitantly increase under these same conditions. Given the robust interaction between mitochondrial MMADHC and the matrix protease LONP1, MMADHC could competitively inhibit LONP1-mediated degradation of TFAM, a key mtDNA nucleoid protein whose abundance correlates with mtDNA copy number.^100–102^ Given MMADHC’s short half-life, it may serve as a LONP1 substrate itself, temporarily sequestering the protease and modulating TFAM turnover. Although complete LONP1 ablation ultimately depletes mtDNA,^103^ subtle modulation through MMADHC levels could provide a physiologically sustainable regulatory mechanism. However, this model remains speculative and requires validation through experiments including direct measurement of TFAM stability upon MMADHC perturbation, biochemical confirmation of LONP1-MMADHC substrate relationships, and functional assessment of whether the MMADHC-LONP1 interaction is necessary for mtDNA regulation.

Due especially to MMADHC’s disease implication, the biological rationale linking a cobalamin pathway protein to mtDNA homeostasis remains an intriguing question warranting further investigation.

Collectively, our work establishes a systematic framework for dissecting mitochondrial proteome regulation and function, providing both a community resource and revealing unexpected biological insights, including the discovery of a novel mitochondrial disease.

Substantial work remains, but these findings represent a significant advance in understanding how mitochondria orchestrate the expression and function of their complex proteome.

### Limitations of the study

The experimental design and technical characteristics lead to some limitations of our study. First, while we profiled 75% of genetic perturbations using two biological replicate clones, we analyzed the remaining knockout lines from single clones, precluding biological replicate validation for these perturbations. Second, filtering analytical findings for congruence across biological replicates was beneficial to remove noise from our results. However, this approach risked excluding relevant signals when replicate clones showed discordant profiles due to inherent genetic and molecular heterogeneity, potentially causing us to miss true biological effects. Third, despite generating broad and deep datasets, most mitochondrial genes were not perturbed, limiting the scope of our findings. Fourth, mitochondrial processes respond flexibly to environmental cues through molecular adaptations. By profiling cell lines exclusively under baseline glucose conditions, we potentially masked relevant biological effects that would emerge under alternative metabolic states or stress conditions.

## METHODS

### Resource availability

#### Lead contact

Requests for further information and resources should be directed to and will be fulfilled by the lead contact, David J. Pagliarini.

#### Materials availability

HAP1 KO cells used in this work were produced in partnership with Horizon Discovery (now part of Revvity, Inc.), or are derivatives thereof, and are subject to their Limited Use Label License. Other cell lines generated in this study can be obtained from the lead contact with a completed material transfer agreement.

### Experimental model and subject details

#### Cell culture

HAP1 wild-type and knockout cell lines (Horizon Discovery) were maintained in IMDM (Thermo 12440053) supplemented with 10% FBS (Sigma F2442) and 1× penicillin-streptomycin (Thermo 15140122). HEK293T and U2OS cells were cultured in DMEM (Gibco 11965-092) with identical supplements. All cells were maintained at 37°C in a humidified incubator with 5% CO_2_.

#### Lentiviral production

We generated lentiviral particles to establish stable cell lines expressing sgRNAs or gene open reading frames. HEK293T LentiX cells (Takara 632180) were seeded in 10 cm dishes 24 hours prior to transfection to achieve 80-90% confluency. Cells were co-transfected with 5 μg of the plasmid of interest along with packaging plasmids pMD2.G (Addgene 12259), pMDLg-pRRE (Addgene 12251), and pRSV-Rev (Addgene 12253) at a 1:4:1 molar ratio using TransIT-Lenti Transfection Reagent (Mirus MIR 6600) in OptiMEM (Gibco 31985-070) according to the manufacturer’s instructions. Forty-eight hours post-transfection, viral supernatants were collected and filtered through 0.45 μm PES filters. We added Lenti-X Concentrator (Takara 631232) at a 1:3 ratio (3.3 mL concentrator per 10 mL supernatant) and incubated the mixture by end-over-end rotation for 1 hour at 4°C. Concentrated virus was pelleted by centrifugation at 1500×g for 45 minutes at 4°C. The viral pellet was resuspended in 500 μL cold PBS, aliquoted in 100 μL portions, flash-frozen in liquid nitrogen, and stored at −80°C. All materials in direct contact with lentivirus were treated with 30% bleach for multiple days before disposal according to biosafety protocols.

### Cell line generation

HAP1 knockout cell lines were generated using CRISPR-Cas9 as previously described.^30^

#### CRISPRi-mediated knockdown

We generated CRISPRi knockdown cell lines using a dual-component system.^104^ Monoclonal HEK293T and U2OS cell lines stably expressing dCas9-KRAB-MeCP2 were established using the lenti_dCas9-KRAB-MeCP2 plasmid (Addgene 122205). For sgRNA expression, we used the pU6-sgRNA EF1Alpha-puro-T2A-BFP plasmid (Addgene 60955). Cells were transduced with lentivirus and selected with 0.5 μg/mL puromycin for 5 days, followed by recovery in antibiotic-free medium for at least 5 days before experiments. The following sgRNAs were used for generation of knockdown lines:

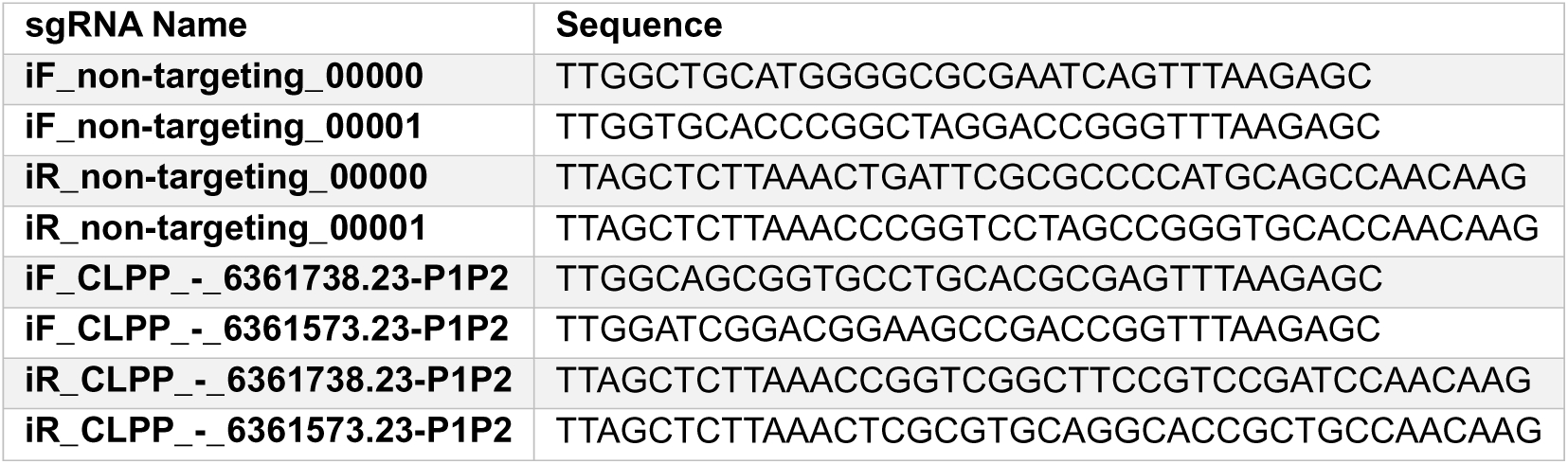

#### CRISPR-Cas9 knockout cell lines

Monoclonal HEK293T cells stably expressing Cas9 were generated using lentiCas9-Blast (Addgene 52962). Knockout cell lines were established by transducing Cas9-expressing cells with lentiGuide-Puro (Addgene 52963) containing target-specific sgRNAs. Following transduction, cells were selected with 0.5 μg/mL puromycin for 5 days and recovered in antibiotic-free medium for at least 5 days before experiments. The following sgRNA sequences were used:

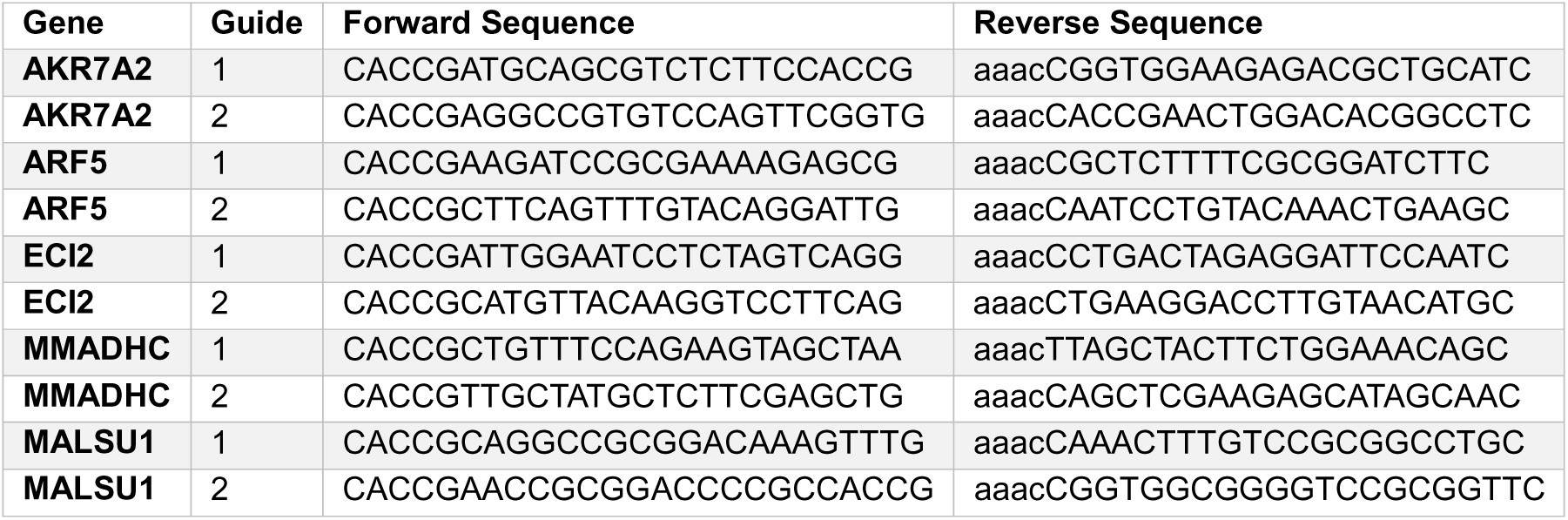

#### Stable rescue and overexpression cell lines

Gene rescue and overexpression cell lines were generated using pLVX-EGFP-IRES-puro (Addgene 128652) or pLV-EF1a-IRES-Hygro (Addgene 85134) vectors containing either EGFP as a control or the gene of interest. The following cDNA sequences were used for overexpression: MALSU1 (NM_138446.2), CLPP (NM_006012.4), MDH2 (NM_005918), ATPAF2 (NM_145691.4), C15orf61 (NM_001143936.2), ATP5ME (NM_007100.4), and MMADHC (NM_015702), or variants thereof. Following lentiviral transduction, cells were selected with 0.5 μg/mL puromycin or hygromycin for 5 days and recovered in antibiotic-free medium for at least 5 days before experiments.

#### Primary fibroblast cell lines

Primary fibroblast samples for MMADHC studies, along with associated biochemical data, were obtained from the University Children’s Hospital Zurich, Switzerland, under ethics approval from the Ethics Committee of the Canton of Zurich (KEK-2014-0211, amendment: PB_2020-00053). Following collection, primary fibroblasts were maintained in Dulbecco’s modified Eagle’s medium (DMEM; Gibco) supplemented with 10% fetal bovine serum (Gibco) and antibiotics (GE Healthcare). After expansion, cells were resuspended in a cryopreservation solution containing 90% fetal bovine serum and 10% dimethyl sulfoxide, then stored in cryovials under liquid nitrogen. For mtDNA measurements, a frozen aliquot of each primary fibroblast line was thawed, washed once with phosphate-buffered saline (PBS), and divided into replicate aliquots.

## Method details

### Bulk RNA-seq

HAP1 cells were cultured in 6-well plates and washed twice with 1 mL cold DPBS (4 °C; Thermo, 14190144) on ice. Cells from three to four biological replicates were lysed in 600 µL Buffer RLT Plus (Qiagen, 1053393) containing 1% β-mercaptoethanol (Sigma, M3148-250ML), mixed by pipetting until homogeneous, transferred to microcentrifuge tubes, and stored at −80°C. Total RNA was purified by Genewiz using the RNeasy Plus Mini Kit (Qiagen). RNA integrity was confirmed prior to Illumina library preparation using PolyA selection (NEBNext Ultra II RNA Library Prep). Sequencing was performed on an Illumina platform with 2 × 150 bp paired-end reads. Base calling and demultiplexing were carried out with Illumina bcl2fastq software, generating raw FASTQ files (yielding a total of 2.5 terabytes of compressed data). RNA-seq reads were aligned to the Ensembl release 101 primary assembly using STAR version 2.7.9a1. Gene-level quantification was performed with Subread:featureCounts version 2.0.3, producing the gene count matrix, while isoform-level quantification of annotated Ensembl transcripts was performed with Salmon version 1.5.23. Sequencing quality was evaluated based on the total number of aligned reads, uniquely mapped reads, and detected features. For differential gene expression profiles, log_2_ fold changes were calculated relative to averaged wild-type controls, and significance was assessed using a two-sided Welch’s *t*-test. The processed RNA-seq data are provided in **Table S8**.

### mtDNA quantification by droplet digital PCR

HAP1 cells grown on 6-well plates were washed twice with 1 mL of cold DPBS (4°C) (Thermo, 14190144) while on ice. Cells were collected by addition of 600 µL RNAprotect Cell Reagent (QIAGEN, 76526), dissociated from plates by pipetting, transferred to a microfuge tube, and stored at −80°C. Absolute quantification of mtDNA genome copy number was performed by Azenta using the QIAcuity Digital PCR platform (Qiagen). Two assays were employed for DNA quantification: B2M_HEX (IDT) with the probe sequence/5HEX/CAGGTTGCT/ZEN/CCACAGGTAGCTCTAG/3IABkFQ/ and ND1_FAM (IDT) with the probe sequence /56-FAM/CCATCACCC/ZEN/TCTACATCACCGCCC/3IABkFQ/. The B2M_HEX assay was performed using 50 ng of DNA input, while the ND1_FAM assay was analyzed at a 1:200 dilution of the 50 ng DNA sample. Whole DNA was extracted using the QIAamp DNA Mini Kit (Qiagen, 51306), and sample quality and concentration were verified with a NanoDrop spectrophotometer. Digital PCR reactions were prepared using the QIAcuity Probe PCR Kit (Qiagen, 250102), following the manufacturer’s quick start protocol. Each 12 µL reaction contained 3 µL of 4x QIAcuity Probe PCR Master Mix, 1.2 µL of 20x probe assay, variable amounts of RNase-free water, and template DNA as indicated. The reactions were loaded onto QIAcuity 8.5k 96-well Nanoplates (Qiagen, 250021) and processed on the QIAcuity dPCR instrument. Thermal cycling was performed under the following conditions: initial heat activation at 95°C for 2 minutes, followed by 40 cycles of 95°C for 15 seconds and 60°C for 30 seconds. Imaging was carried out using multiple fluorescent channels with the following exposure and gain settings: FAM (500 ms, gain 6), HEX (500 ms, gain 6). Data analysis was performed using QIAcuity Software Suite v2.2.0.26. Each run included a no-template control (NTC) to confirm the absence of contamination. Thresholds distinguishing positive and negative partitions were manually set in the 1D scatterplot to ensure consistency across runs. ND1 copy numbers were corrected for the 200-fold dilution and normalized to the corresponding B2M copy number values. The resulting ND1/B2M ratios represent the mitochondrial DNA copy number, which was further normalized to wild-type samples to calculate log_₂_ fold-changes and assess statistical significance using a two-sided Welch’s *t*-test.

### Isolation of crude mitochondria

Unless mentioned otherwise mitochondrial isolates were obtained from cultured cells following a standard differential centrifugation protocol designed to preserve organelle integrity. Several days before isolation, cells were plated and grown to approximately 90% confluency. On the day of harvest, culture media was removed, and cells were washed once with DPBS. They were then detached using a cell scraper and fresh ice-cold PBS and transferred to prechilled 50 mL conical tubes kept on ice. After collecting all plates of the same sample group (*e.g.*, cell line), the cell suspension was centrifuged at 600×g for 10 min at 4°C, washed once with 10 mL ice-cold PBS, and pelleted again at 600×g for 5 min. The resulting pellet was resuspended in 3 mL ice-cold mitochondrial isolation buffer (210 mM mannitol (Sigma, M4125), 70 mM sucrose (Sigma, S9378), 5 mM HEPES pH 7.4 (RPI, H75030), 1 mM EGTA (Sigma, E4378) freshly supplemented with protease inhibitors (Roche, Complete EDTA-free, 05056489001). A separate aliquot of the same buffer was prepared containing 0.5% fatty acid–free BSA (Sigma, A7030) and used exclusively for resuspension and homogenization of cells. Cells were transferred to a prechilled 7 mL Dounce homogenizer (DWK Wheaton, EW-04468-02) and disrupted on ice with 30–35 strokes using a tight pestle. The homogenate was centrifuged at 600×g for 10 min at 4°C to remove nuclei and debris. The supernatant, containing mitochondria and cytosolic components, was transferred to fresh microcentrifuge tubes and spun at 7000×g for 10 min at 4°C. The resulting pellet represented the mitochondria-enriched fraction, while the supernatant corresponded to the cytosolic fraction. The mitochondrial pellet was washed once with 200 µL prechilled isolation buffer and centrifuged again at 7000×g for 10 min at 4°C. The final pellet was gently resuspended in 200 µL mitochondrial isolation buffer using wide-bore tips to avoid mechanical damage. Protein content was determined by a Pierce BCA Protein Assay Kit (Thermo, 23225) and mitochondria were aliquoted. The resulting preparation constituted the crude mitochondrial fraction, which was used immediately for downstream experiments or flash-frozen and stored at −80°C for later use. If mitochondrial pellets were flash-frozen, aliquoted mitochondria was centrifuged again at 7000×g for 10 min at 4°C and all the supernatant removed.

### Complexome profiling

#### Sample preparation

Crude mitochondria were resuspended in solubilization buffer (50 mM imidazole (Thermo, 122020020), 500 mM 6-aminocaproic acid (Sigma, A2504), 1 mM EDTA (Sigma, 03690) at 10 µL per 100 µg of mitochondrial protein content. Membranes were homogenized gently by twirling a small spatula, and digitonin (Millipore Sigma, D141-500MG) was added to a final detergent-to-protein ratio of 6 g/g. Samples were incubated on ice for 20 min to solubilize, followed by centrifugation at 20,000×g for 20 min at 4°C to remove insoluble material. The supernatant containing solubilized complexes was transferred to a clean tube, and protein concentration was quantified using a Pierce BCA Protein Assay Kit (Thermo, 23225). The sample was then supplemented with 50% glycerol to a final concentration of 5% (w/v) and Coomassie Brilliant Blue G-250 (VWR, M140-50G) to a final detergent-to-dye ratio of 8 g/g, ensuring protein stability and optimal visualization for Blue Native PAGE.

#### Blue native PAGE

Protein complexes were separated using a Bio-Rad Protean Cell II xi system according to the manufacturer’s instructions. Self-cast 4–13% large-format native gels (1.5 mm x 14 cm (height) x 15 wells (width)) were prepared as described.^105^ Wells were rinsed with BN Anode Buffer (25 mM imidazole, ThermoFisher 122020020, pH 7.0) before loading 50 µg of lysate per well and 10 µL of unstained NativeMark protein ladder (ThermoFisher, LC0725). The outer chamber was filled with BN Anode Buffer, and the inner chamber with BN Cathode Buffer B (50 mM tricine (Sigma, T0377), 7.5 mM imidazole, 0.02% Coomassie Blue G-250). Electrophoresis was run at 4°C at 100 V constant voltage until the sample entered the gel (∼1 h), then continued at 15 mA constant current. Once the dye front migrated one-third of the gel, Buffer B was replaced with Buffer B/10 (50 mM tricine, 7.5 mM imidazole, 0.002% Coomassie Blue G-250), and the run continued until the dye front reached the bottom (∼3-4 h). Gels were then removed, stained with Coomassie, and destained repeatedly before incubation in water overnight. Each lane was subsequently sliced into 60 equal gel fractions for downstream analysis.

#### In-gel tryptic digest

Each gel slice was cut into small cubes, transferred to a 96-well filter plate, and destained with 100 µL destaining buffer (50% LC/MS-grade acetonitrile, 50 mM ammonium bicarbonate) with gentle agitation. After five washes, reduction and alkylation were performed using Tris(2-carboxyethyl)phosphine (100 mM final, sigma aldrich C4706-10G) and 2-chloroacetamide (400 mM final, sigma aldrich C0267) in 25 mM ammonium bicarbonate (sigma aldrich A6141-500G) for 15 min at room temperature. Gel pieces were dehydrated twice with acetonitrile (fisher chemical A955-4), air-dried, and rehydrated with trypsin (Promega Sequencing Grade, 0.4 µg/µL; diluted 1:25 to ∼16 ng/µL final) in 25 mM ammonium bicarbonate. Samples were incubated 15 min at room temperature, then more digestion buffer (25 mM ammonium bicarbonate) was added, and digestion was performed at room temperature overnight under gentle agitation. The next day, peptides were retrieved by centrifugation and additionally extracted using 100 µL extraction buffer (50% acetonitrile, 5% formic acid (Thermo, 28905). The sample (∼200 µL total volume) was dried by SpeedVac at 40°C. The dried material was resuspended in 0.2% TFA (Thermo, 85183), briefly sonicated, and desalted using a Peptide Clean-up Plate (Thermo, A57865). After confirming sample acidity (pH < 2), peptides were loaded onto the desalting plate, washed sequentially with 5% acetonitrile/0.2% formic acid and 100% acetonitrile, and eluted with 40% acetonitrile/5% ammonium hydroxide. Eluates were dried at 40°C, resuspended in 50 µL 0.2% formic acid, and briefly sonicated before LC-MS analysis.

#### Liquid chromatography method

Peptide separation was performed using a Vanquish Neo UHPLC system (Thermo Scientific) operated under a 12.8-minute LC gradient. A 1 µL injection volume was loaded at a flow rate of 100 µL/min onto a trap column (300 µm × 0.5 cm, backward flush mode) and subsequently eluted onto a µPAC HT Neo HPLC analytical column (Thermo COL-CAPHTNEOB, 75 µm × 5.5 cm) maintained at 50°C. The system operated in Trap-and-Elute mode within the NanoCap flow regime, using water (solvent A) and 80% acetonitrile (solvent B) as mobile phases. The column flow rate was kept constant at 1.5 µL/min, and pressure was limited to 450 bar. The LC gradient began at 1.2% B at 0.0 min, increased to 10% B by 0.1 min, then gradually to 28% B by 8.1 min and 56% B by 11.7 min. The column was then rapidly washed to 99% B at 11.82 min, maintained for column cleaning until 12.8 min, and followed by automated re-equilibration. Fast column equilibration (flow = 3 µL/min; pressure = 450 bar) and multi-step trap washes (four cycles, 800 bar) were enabled to ensure reproducibility. The autosampler temperature was maintained at 7 °C, and injection parameters included a draw speed of 0.2 µL/s, dispense speed of 5 µL/s, and sequential weak and strong needle washes (5 s and 3 s, respectively).

Peptides were separated on the µPAC column under these conditions using Thermo Scientific SII for Xcalibur software (version 4.7.69.37). Samples were run sequentially from the lowest to highest molecular weight gel slice (slices 1–60).

#### Mass spectrometry data acquisition

Mass spectrometric data were acquired on an Orbitrap Astral Mass Spectrometer (Thermo Fisher Scientific) operated in data-independent acquisition (DIA) mode. The ion source operated under standard LC conditions without FAIMS, using positive polarity and applying an RF lens voltage of 40%. Lock mass correction was disabled, and advanced peak determination was enabled to enhance precursor feature detection. The system was configured for an expected LC peak width of 6 seconds and a default charge state of +2. The acquisition method comprised two sequential experiments. The first (Experiment 1: MS1 full scan) was performed in the Orbitrap detector at a resolution of 240,000 (at m/z 200) across a scan range of 380–980 m/z.

The AGC target was set to 3.0 x 10L (300% normalized), with a maximum injection time of 5 ms and one microscan per spectrum. Data were collected in profile mode, with source fragmentation disabled. The second (Experiment 2: DIA) was performed in Astral detection mode across the same precursor mass range (380–980 m/z) using automatically defined 2 Th isolation windows with 0 Th overlap, resulting in 299 variable scan events. Window placement optimization was enabled to maximize spectral coverage. Fragmentation was achieved using higher-energy collisional dissociation (HCD) at a normalized collision energy of 26%. MS2 scans were recorded in the m/z 150–2000 range, in centroid mode, with an AGC target of 5.0 x 10L (500% normalized) and a maximum injection time of 3 ms. The DIA cycle was operated in time-based loop control mode with 0.6-second cycle times, ensuring comprehensive sampling across chromatographic peaks. All data were acquired using Thermo Scientific Orbitrap Astral Control Software in direct synchronization with the LC gradient.

### Proteomics of HAP1 KO cell collection

The proteomics data based on the HAP1 KO cell collection was used as previously published^30^ with two minor modifications to ensure unambiguous protein naming and up to date mitochondrial localization nomenclature. First, empty HGNC symbols were replaced with the UniProt protein ID, then 171 duplicate entries were removed while retaining the duplicate with the overall higher magnitude of log_2_ fold changes (8433 proteins previously, now 8262). Second, the annotation for mitochondrial protein localization was updated to MitoCarta3.0^17^ from MitoCarta2.0. The proteomics data table used in this study is provided in **Table S9**.

### Merged transcriptomics-proteomics data

The updated proteomics dataset and the newly obtained RNA-seq datasets were merged based on gene identifiers to enable unique matches. This merged data table is provided as **Table S10**.

### Proteomics sample processing

In addition to the previously published proteomics dataset and the complexome profiling dataset described in this methods section, a number of further proteomics experiments were performed in this study.

#### Sample preparation

Either whole-cell lysates or crude mitochondrial fractions were used as starting material. Pellets stored at −80 °C were thawed on ice and resuspended in 150 µL lysis buffer (8 M urea, 100 mM Tris-HCl pH 8.0, 1× HALT protease/phosphatase inhibitor, Thermo, 78440). Samples were sonicated using a probe sonicator (1 × 10 seconds pulse, 10% output) and clarified by centrifugation at 16,500×g for 15 min at 4 °C. Protein concentrations were determined using the BCA assay (Pierce, Thermo, 23225). Approximately 100 µg protein from each sample was transferred to a low-protein-binding microcentrifuge tube. Samples were reduced by addition of tris(2-carboxyethyl)phosphine to a final concentration of 10 mM and alkylated with 2-chloroacetamide to a final concentration of 40 mM. The volume was adjusted to 150 µL with 100 mM Tris-HCl pH 8.0, and samples were incubated for 1 h at room temperature (1000 rpm) for reduction and alkylation. Proteins were digested with trypsin (Promega, V5113 or Thermo, 90305) at a 1:50 enzyme-to-protein ratio, ensuring a final urea concentration below 2 M. CaCl_₂_ was added to a final concentration of 1 µM, and the reaction volume was adjusted with 100 mM Tris-HCl pH 8.0. Digestion proceeded overnight (16–20 h) at 37°C and 1000 rpm in a thermomixer.

#### Desalting of peptides

For peptide cleanup prior to LC–MS analysis on the **Orbitrap Astral**, peptides were desalted using Pierce Peptide Desalting Spin Columns (Thermo, 89851) following the manufacturer’s instructions with minor modifications. Columns were activated twice with 300 µL ACN and equilibrated twice with 300 µL 0.1% TFA by centrifugation at 5,000×g for 1 min. Acidified peptide samples were loaded, centrifuged at 3,000×g for 1 min, and washed twice with 300 µL 0.1% TFA. Peptides were eluted twice with 300 µL 50% ACN/0.1% TFA into fresh 2 mL LoBind tubes, yielding a total elution volume of 600 µL. For analysis on the **Exploris 240**, Strata X columns (Phenomenex, 8B-S100-AAK) were prepared on the day of use. Buffers included 0.1% trifluoroacetic acid (TFA) in water and 80% acetonitrile (ACN) with 0.1% TFA. Columns were conditioned sequentially with 1 mL 100% ACN followed by 1 mL 0.1% TFA. Samples were acidified with a few µL of 50% TFA, checked by pH paper to confirm acidity, and centrifuged at 16,000×g for 5 min at 4°C. Supernatants were loaded onto the columns, washed once with 1 mL 0.1% TFA, and peptides were eluted with 300 µL 80% ACN/0.1% TFA into low-protein-binding tubes.

In both workflows, eluates were dried in a SpeedVac. Then dried peptides were resuspended in 0.2% formic acid immediately prior to injection or stored at −80 °C for later use.

#### Peptide measurement and injection

Peptide concentrations were quantified after resuspension in 0.2% formic acid using the NanoDrop OneC spectrophotometer (Thermo, 31310) with Protein A205 method 31, or alternatively by fluorescence-based quantification (Thermo, 23290) according to the manufacturer’s protocol. For LC–MS acquisition, 1 µg peptide was injected for analysis on the Exploris 240 and 200 ng for analysis on the Orbitrap Astral.

### Proteomics data acquisition

Proteomics experiments were performed on two different instruments (Orbitrap Exploris 240 and Astral mass spectrometers by Thermo Scientific), necessitating two designated workflows.

#### Liquid chromatography method

For subsequent analysis on the **Orbitrap Astral** mass spectrometer, peptide separation was performed using a Vanquish Neo UHPLC system (Thermo Scientific) operated under a 31-minute LC gradient. A 1 µL injection volume was loaded at a flow rate of 20 µL/min onto a trap column (300 µm × 0.5 cm, backward flush mode) and subsequently eluted onto an Aurora Ultimate 25 × 150 XT C18 UHPLC analytical column (IonOpticks AUR4-250150C18-XT) maintained at 40°C. The system operated in Trap-and-Elute mode within the NanoCap flow regime, using water with 0.1% formic acid (solvent A) and 80% acetonitrile with 0.1% formic acid (solvent B) as mobile phases. The analytical flow rate was set to 0.5 µL/min with a pressure limit of 1500 bar. The LC gradient started at 4% B and was held until 0.5 min, increased to 8% B by 0.6 min, then gradually to 35% B at 25.8 min and 55% B at 26.2 min, followed by a rapid ramp to 99% B at 26.7 min for column washing, which was maintained until 31.0 min before re-equilibration. Fast column equilibration (0.8 µL/min, 400 bar) and two automated trap wash cycles at 800 bar were enabled to ensure reproducibility. The autosampler temperature was maintained at 7°C, with injection parameters including a draw speed of 0.2 µL/s, dispense speed of 5 µL/s, and sequential weak and strong needle washes of 5 s and 3 s, respectively. Peptides were separated under these conditions using Thermo Scientific SII for Xcalibur software (version 4.7.69.37).

For subsequent analysis on the **Orbitrap Exploris 240** mass spectrometer, peptide separation was performed on an RSLC-nano system (Thermo Scientific) using a 120-minute run method. The autosampler operated in low-dispersion pick-up mode with a nominal wash volume of 50 µL, draw speed 0.2 µL/s, dispense speed 2 µL/s, a dispense delay of 2 s and a controlled wash speed of 4 µL/s; the sampler temperature was maintained at 6°C. Loading pump conditions were set to deliver 5.0 µL/min during sample loading, with the loading pump solvent composition defined as 98% water / 2% ACN / 0.1% TFA and a permitted pressure window up to 500 bar. An EASY-Spray HPLC column (Thermo ES902, 75 µm × 250 mm) analytical column was used and maintained at ambient temperature. The nanoflow pump used 0.1% formic acid in water as solvent A and 0.1% formic acid in 80:20 ACN:water as solvent B, with a nominal analytical flow of 0.300 µL/min and a maximum allowable pressure of 800 bar. The programmed gradient began with the NC pump at 4% B at 3.0 min, progressed to 20% B at 60.0 min, to 45% B at 100.0 min, and was ramped to 99% B at 105.0–110.0 min for column washing, returning to 4% B by 111.0 min for re-equilibration through the end of the 120-min method. Rapid flow ramping limits and controlled ramp profiles were applied (NC_Pump flow ramp ±0.300 µL/min^2^; LoadingPump ramp ±5 µL/min^2^) and two automated trap wash cycles were enabled. All steps were executed under the instrument method using Thermo Scientific SII for Xcalibur (version 4.6.67.17).

#### Mass spectrometry data acquisition

For acquisition of mass spectrometric data on the **Orbitrap Astral** mass spectrometer (Thermo Fisher Scientific) the instrument was operated in data-independent acquisition (DIA) mode over a total run time of 31 minutes. The ion source was a nanospray ionization source operated in static mode at a spray voltage of +1500 V under positive polarity, with the ion transfer tube maintained at 280°C. No FAIMS interface was used. EASY-IC internal calibration was applied at run start for lock mass correction. The acquisition consisted of two sequential experiments. The first experiment (MS1 full scan) was performed in the Orbitrap detector at a resolution of 240,000 (at m/z 200) over a scan range of 380–980 m/z. The RF lens voltage was set to 40%, with the AGC target defined at 5 × 10^6^ (500% normalized) and a maximum injection time of 3 ms. One microscan was acquired per spectrum in profile mode, with source fragmentation disabled. The second experiment (DIA) covered the same precursor mass range (380–980 m/z) using 2 m/z isolation windows without overlap, resulting in 300 scan events. Window placement optimization was disabled, and scans were acquired in Astral detection mode over an m/z range of 150–2000. Fragmentation was achieved by higher-energy collisional dissociation (HCD) with a normalized collision energy of 25%. MS2 spectra were collected in centroid mode with an AGC target of 5 × 10^4^ (500% normalized), a maximum injection time of 3 ms, and one microscan per spectrum. The DIA cycle operated in time-based loop control with a 0.6-second cycle time. Advanced peak determination was enabled, with an expected chromatographic peak width of 6 seconds and a default precursor charge state of +2. Data acquisition was performed using Thermo Scientific Orbitrap Astral Control Software in synchronization with the LC gradient.

For acquisition of mass spectrometric data on the **Orbitrap Exploris 240** mass spectrometer (Thermo Fisher Scientific) the instrument was operated in data-dependent acquisition (DDA) mode over a total run time of 120 min, using a nanospray ionization source in static mode (positive polarity) with a spray voltage of +1800 V, negative ion voltage set to −600 V, and the ion transfer tube maintained at 275 °C; a carrier gas flow of 1.2 L/min was applied at the start of the run. Internal mass calibration (EASY-IC) was enabled at run start. Global acquisition parameters specified an expected chromatographic peak width of 20 s and enabled advanced peak determination with a default precursor charge state of +2. The method began with a full MS survey scan in the Orbitrap (resolution 120,000) across m/z 375–1,500, RF lens 80%, with custom AGC (normalized target 300%) and a maximum injection time of 50 ms (one microscan), acquired in profile mode. Precursor selection employed monoisotopic peak determination tuned for peptides and an intensity threshold of 1.0×10^5^, considering charge states +2 to +6; isotope peaks were excluded. Dynamic exclusion removed a precursor after one selection for 60 s with a mass tolerance of ±10 ppm. Data-dependent acquisition ran in cycle-time mode with 3 s between master scans; dependent MS2 events were performed as ddMS^2^ with an isolation window of 1.6 m/z, HCD collision energy 25% (normalized), MS2 detection in the Orbitrap at 30,000 resolution (first mass m/z 110), AGC normalized target 75%, maximum injection time set to automatic, and MS2 spectra recorded in centroid mode.

### Crosslinking affinity enrichment proteomics (XL-AEMS)

Crosslinking affinity enrichment proteomics experiments were conducted using either whole-cell lysates or crude mitochondria as starting material. Baits were enriched via FLAG antibody pulldown (for FLAG-tagged constructs introduced permanently via transduction or transiently via transfection) or by immunoprecipitation with antibodies against the endogenous protein. The endogenous approach was exclusively performed using crude mitochondria and included corresponding knockout (KO) lines as negative controls.

#### Crosslinking of proteins

Protein crosslinking was carried out using disuccinimidyl sulfoxide (DSSO), a cleavable crosslinker that covalently links proximal primary amine groups (typically those of lysine residues) via stable amide bonds. For crosslinking of **intact mitochondria**, 500–1000 µg of crude mitochondria was thawed on ice, centrifuged at 15,000×g for 5 min at 4°C, and the supernatant was aspirated. Each pellet was gently resuspended in HEENK buffer (10 mM HEPES, pH 7.1, 1 mM EDTA, 1 mM EGTA, 10 mM NaCl, 150 mM KCl) using wide-bore tips, typically adding 100 µL for a pellet of approximately 100 µL to achieve a final volume of around 200 µL once DSSO was added. Crosslinking was performed by addition of DSSO to a final concentration of 0.5 mM (Thermo A33545; 50 mM stock prepared by dissolving 1 mg DSSO in 51.5 µL DMSO) and incubation for 1 h at room temperature without shaking. The reaction was quenched with Tris, pH 8.0, to a final concentration of 100 mM. Mitochondria were pelleted by centrifugation (15,000×g, 5 min, 4°C), washed once with 1 mL HEENK containing 100 mM Tris without resuspension, and centrifuged again under the same conditions.

For DSSO crosslinking of **intact cells**, cells were seeded in 10 cm dishes one day prior to crosslinking to reach approximately 70 % confluence at the time of treatment. Transfection was performed using Lipofectamine 3000 (Invitrogen L3000075), and 24 h post-transfection, cells were washed once with PBS and collected using a cell scraper. Pelleted cells were resuspended in 1 mL PBS containing 2 mM DSSO (40 µL of 50 mM stock) and incubated for 1 h at 37°C with shaking at 200 rpm to allow crosslink formation. Pellet cells (1000×g, 3 min, 4°C) and wash twice with 1 mL of 50 mM Tris-HCl (pH 8.0) to quench the reaction. Pellets were frozen at –80°C or used for immediate processing.

#### Immunoprecipitation

For **FLAG immunoprecipitation from intact mitochondria**, crosslinked mitochondria were solubilized in 50-100 µL solubilization buffer (50 mM imidazole, 500 mM 6-aminocaproic acid, 1 mM EDTA, supplemented immediately before use with 1× Halt protease/phosphatase inhibitor) containing digitonin at a ratio of 6 g per g protein (15-30 µL of 20% digitonin stock). Pellets were gently resuspended using a small spatula to preserve protein complexes and incubated on ice for 20 min. Insoluble material was removed by centrifugation at 15,000×g for 20 min at 4°C. The supernatant was added to pre-washed Magnetic Anti-FLAG M2 Beads (Sigma M8823-1ML; 30 µL per sample), which had been washed three times with Low Salt Wash Buffer (LSWB; 300 mM NaCl, 20 mM HEPES pH 7.8, 20 % glycerol, 2 mM MgCl_2_, 0.2 mM EDTA, 0.05 % digitonin, 0.5 mM DTT, and protease inhibitors). The mixture was incubated with rotation at 4°C for 1 h, adjusting the total volume to at least 200 µL with solubilization buffer. Beads were subsequently washed twice each with LSWB, PBS + 0.05 % digitonin, and PBS (total of six washes using 500 uL buffer, incubate for 2 min at 4°C).

For **endogenous protein immunoprecipitation from intact mitochondria**, solubilization was performed as described above. The supernatant was adjusted to a total volume of 250 µL with LSWB and incubated overnight at 4°C with either rabbit IgG or an antibody against the bait protein (1.5 µg of antibody per sample) under end-over-end agitation. The antibodies used were MIC60/IMMT (Proteintech 10179-1-AP), MDH2 (Abcam ab181873; clone EPR14882(B)), and rabbit IgG control (Thermo 02-6102). Each experiment included four conditions: WT + IgG, WT + bait Ab, KO + IgG, and KO + bait Ab (performed in triplicate). The next day, 30 µL Protein A beads (Thermo 10001D) per IP were washed once with water and twice with LSWB before addition of the lysates. After 2 h of incubation at 4 °C with rotation, beads were washed twice with LSWB, PBS + 0.05 % digitonin, and PBS (same as above).

**FLAG immunoprecipitation from intact cells** was carried out in a similar manner. Magnetic Anti-FLAG M2 Beads (30 µL per sample) were washed three times with LSWB containing 300 mM NaCl, 20 mM HEPES pH 7.8, 20 % glycerol, 2 mM MgCl_₂_, 0.2 mM EDTA, 1 % NP-40, 0.5 mM DTT, and protease inhibitors. Cross-linked cell pellets were resuspended in 500 µL lysis buffer (150 mM NaCl, 20 mM HEPES (pH 7.8), 20% glycerol, 2 mM MgCl_2_, 1 mM EDTA, 1% NP-40 (IGEPAL), 1 mM DTT, 1x EDTA-free protease inhibitor), vortexed and incubated on ice for 10 min. Lysates were cleared by centrifugation (15,000×g, 10 min, 4°C) and the protein content was determined using a BCA assay (Thermo, 23225). 1-2 mg of total protein (same amount per sample) were added to the equilibrated beads and rotated at 4°C for 1 h, ensuring a total volume of at least 500 µL. Beads were then washed twice each with LSWB, PBS + 1 % NP-40, and PBS (same as above).

#### On-bead trypsin digest

Following immunoprecipitation, all samples were processed identically. On-bead proteins were denatured in 200 µL 2 M urea in 100 mM Tris pH 8.0, reduced with 5 mM DTT at 56°C for 30 min with shaking at 1000 rpm, and alkylated with 15 mM iodoacetamide for 30 min at room temperature in the dark. The reaction was quenched with an additional 1 µL of 1 M DTT, and samples were digested overnight at 37°C with 1 µg trypsin at 1000 rpm. The following day, supernatants were transferred to new tubes, beads were washed with 200 µL 200 mM ammonium bicarbonate, and both fractions were combined. The pH was adjusted to below 2 by addition of a few microliters of 50% TFA. Peptides were desalted using Pierce Peptide Desalting Spin Columns (Thermo 89851) according to the manufacturer’s instructions, dried by vacuum centrifugation, and stored at −80°C. Prior to LC–MS analysis, peptides were resuspended in 0.2% formic acid, and equal volumes per sample were injected, targeting a total ion current (TIC) value of approximately 1 × 10^9^ for the most abundant sample.

### Sucrose gradient mitoribosome sedimentation

#### Preparation of continuous sucrose gradient by diffusion

Continuous sucrose gradients were prepared using methanol-washed 5 mL Open-Top Thinwall Polypropylene Tubes (13 x 51 mm; Beckman Coulter, 326819). The midline of each tube was marked to define the approximate midpoint of the gradient. Two sucrose solutions were prepared in Tris-NaCl-MgCl_₂_ buffer (TNM: 50 mM Tris-HCl, pH 7.4; 100 mM NaCl; 20 mM MgCl_₂_) containing either 7.5% (w/v) or 30% (w/v) sucrose, each adjusted to 10 mL total volume with ddH_₂_O and kept on ice. Each ultracentrifuge tube was first filled to the mark with 7.5% sucrose-TNM, after which ∼2.5 mL of the 30% sucrose-TNM solution was gently loaded beneath it using a syringe fitted with a blunt needle, inserting the needle to the bottom of the tube and dispensing slowly until the interphase reached the marked midpoint. The tubes were sealed with parafilm, placed securely in a rack, and the rack was gradually tilted onto its side over 30 seconds, allowing the layers to diffuse horizontally overnight at 4°C without agitation. The following day, the rack was slowly returned to the upright position over another 30 seconds. The resulting gradient was used for sedimentation experiments.

#### Sample lysis and ultracentrifugation

Mitochondrial pellets were resuspended in 250 µL of freshly prepared lysis buffer (50 mM Tris-HCl (Merck 648313), pH 7.4; 150 mM NaCl (Sigma S7653); 1 mM EDTA; 1% (v/v) Triton X-100; 1x protease inhibitor cocktail (Thermo Scientific 1861281); 1 U/mL RNase inhibitor (Promega N211A); ddH_₂_O to 50 mL total volume). The buffer was filter-sterilized, aliquoted, and stored at 4°C, with RNase inhibitor added immediately before use. Samples were incubated on a roller at 4°C for 15–20 min to ensure efficient lysis, then clarified by centrifugation at 13,000×g for 5 min at 4°C. Preformed 7.5-30% continuous sucrose gradients were removed from 4°C immediately before use and handled gently to avoid disturbing the gradient. From each gradient tube, 200 µL of liquid was removed from the top, and 200 µL of clarified mitochondrial lysate was carefully loaded by dispensing against the inner wall of the tube. Approximately 50 µL of residual lysate was retained and frozen at −20°C as a loading control. Gradients were centrifuged in a Beckman Coulter SW55Ti rotor at 40,000 rpm (average RCF = 151,693×g) for 3 h 15 min at 4°C. Following centrifugation, gradients were manually fractionated from top to bottom in 400 µL increments into clean 2 mL microcentrifuge tubes. Fraction 1 corresponded to the top (lowest sucrose concentration, smallest particles) and subsequent fractions represented progressively denser sucrose layers toward the bottom.

#### TCA precipitation of fractions

Each collected fraction (400 µL) was diluted 1:3 with sucrose-free gradient buffer (50 mM Tris-HCl, pH 7.4; 100 mM NaCl; 20 mM MgCl_2_ (Sigma M8266)) to a final volume of 1.6 mL. One-quarter volume of 100% trichloroacetic acid (Sigma, T9159) was added, bringing the total volume to 2 mL. Samples were vortexed briefly and incubated overnight at −20°C to precipitate proteins. The next day, samples were thawed at room temperature for 15–30 min, vortexed, and centrifuged at 14,000 rpm for 15 min at 4°C. The supernatant was carefully aspirated, and pellets were washed once with 2 mL of ice-cold 100% acetone (Fisher Scientific, A18-4). Tubes were centrifuged again at 14,000 rpm for 10 min at 4°C, the supernatant was removed, and the pellets were air-dried for 5–10 min at room temperature to evaporate residual acetone. Once dry, tubes were sealed and stored at –20 °C until further use.

#### Western blotting of fractions

Protein pellets from each fraction were resuspended in 70 µL of 2x Laemmli sample buffer supplemented with DTT (1:10 dilution) and boiled for 5 min at 95°C. For the mitochondrial lysate loading control, 22 µL of lysate was mixed with 58 µL of 2x Laemmli buffer containing DTT and boiled for 5 min at 95°C. A total of 18 µL of each fraction was loaded per lane onto a 15-well 10% Bis-Tris gel, providing sufficient material for up to three replicate gels (3 x 18 µL). Eighteen microliters of the mitochondrial lysate loading control was loaded in the first lane of each gel. Subsequent steps were performed according to the standard Western blotting protocol.

### Sucrose gradient SILAC proteomics

#### Protein labeling in cell culture

To allow quantitative comparison of cell proteomes in sucrose gradient fractions between wild-type and CLPP-KO genotypes, cells were grown in the presence of amino acids containing different stable isotopes until most of their proteins have incorporated them. HAP1 cells (wild-type and CLPP-KO) were cultured in IMDM for SILAC media (thermo scientific 88367). 56.5 mL of media have been removed from a 500 ml bottle, which then has been supplemented with dialyzed FBS (50 ml), Penicillin/Streptomycin (5ml), L-proline (500 uL of 200 mg/mL stock, Sigma P0380), Arg, medium or heavy (500 uL of 100 mg/mL stock, Cambridge Isotope Laboratories Inc. CLM-2265-H-PK or CNLM-539-H-PK), Lysine (500 uL of 100 mg/mL stock, Cambridge Isotope Laboratories Inc. DLM-2640-PK or CNLM-291-H-PK). We started culturing WT cells with medium and CLPP-KO cells with heavy SILAC medium. Cells were maintained in culture in 10 cm dishes with these media and after 10 days of culturing the label incorporation was >97% as detected by mass spectrometry. Then cells were split and expanded over the following days to yield ten 15 cm dishes for all four conditions.

#### Harvesting cells and determining protein concentration

Cells were harvested after reaching confluency for isolation of crude mitochondria. Remove the SILAC media from the cells. Add 5-10 ml ice-cold PBS to wash the cells. Remove PBS and add 2 ml of fresh ice-cold PBS to each plate. Detach cells using a cell lifter (Corning Incorporated 3008). Transfer cell suspension into a 50 ml conical on ice. Once all cells have been collected, centrifuge at 500×g at 4°C, 4 minutes. Discard the supernatant. Resuspend in 10 mL ice-cold PBS, spin 500×g 5 min, discard supernatant. Resuspend in 10 mL ice-cold PBS. Cell suspension is kept on ice. Remove 20 ul of each suspension (medium and heavy) for each cell line into a new 1.5 mL centrifuge tube. Pellet the cells at 600×g at 4°C for 3 minutes and carefully remove the supernatant. Resuspend each pellet in 20 uL lysis buffer (Tris-HCl pH 7.4, 150 mM NaCl, 1mM EDTA, Triton X-100 1%, protease inhibitor 1×). Incubate on ice for 5 minutes. Centrifuge at 16,500×g, 4°C for 5 min and transfer supernatant into a new microcentrifuge tube. Perform BCA and combine samples from the two conditions according to the relative factor that is calculated based on the BCA.

#### Sucrose gradient and proteomics sample preparation

Continuous sucrose gradient preparation, mitochondrial sample lysis, and ultracentrifugation were performed as described above. For this experiment 25 fractions were obtained (200 µL each) and the protein was precipitated using TCA as described above. Precipitated samples were resuspended in 50 µL lysis buffer (8 M urea, 100 mM Tris-HCl pH 8.0, 1× HALT protease/phosphatase inhibitor, Thermo, 78440). No sonication was performed and reduction/alkylation as well as trypsin digest was performed as described in the proteomics section above. After trypsin digest overnight, samples were desalted using the Strata X columns as described above and dried samples were resuspended in 30 µL 0.2% formic acid and 2 µL of sample were injected on the Orbitrap Exploris 240.

### Transient protein overexpression

We seeded 3.5×10^5^ wild-type HEK293T cells per well in poly-L-lysine-coated 6-well plates with DMEM supplemented with FBS and penicillin/streptomycin. Twenty-four hours post-seeding, we switched cells to glucose-free DMEM containing 10 mM galactose, dialyzed FBS, penicillin/streptomycin, and 1× GlutaMAX (Gibco, 35050061). Cells reached ∼70% confluency after one day and we replaced media with 2 mL fresh galactose DMEM per well and transfected cells using Lipofectamine 3000 (Invitrogen, L3000015) with 2 μg plasmid DNA according to the manufacturer’s protocol. We transfected the following plasmids: COQ8A (NM_020247.5), COQ8B (NM_024876.4), COQ8A+COQ8B, or EGFP (control). Forty-eight hours post-transfection, we harvested cells, froze pellets at −80°C, and processed samples later for proteomics analysis as described above.

### CLPP activation with ONC201

To pharmacologically activate ClpP, U2OS CRISPRi control (non-targeting) and CLPP knockdown cell lines were treated with the ClpP agonist ONC201 (TIC10; SelleckChem, S7963). The compound stock solution is concentrated at 10 mM in DMSO and was stored at −20°C. Cells were seeded in 6-well plates at a density of 8 × 10LJ cells per well in DMEM with standard supplements and cultured overnight before treatment. The following conditions were applied in duplicate for each cell line: vehicle control (0.1% DMSO), and ONC201 at final concentrations of 0.1 µM, 1 µM, and 10 µM (each maintained at a consistent solvent concentration of 0.1% DMSO). After compound addition, cells were incubated under standard conditions for 24 hours, then harvested by centrifugation, washed once with cold PBS, and frozen at −80 °C until analysis. Protein levels were evaluated by Western blotting.

### Cycloheximide treatment

Cycloheximide solution (Sigma, C4859) was used at a 100 mg/mL stock concentration in dimethyl sulfoxide (DMSO). HAP1 wild-type and CLPP knockout cells were seeded in one 10 cm dishes per condition at a density of 3 × 10^6^ cells per well and cultured to 70% confluency the following day. At time zero, the first set of plates was washed once with pre-warmed DPBS and immediately harvested to serve as the baseline control. Cycloheximide treatment medium was prepared by diluting the 100 mg/mL stock 1:2000 in complete IMDM to achieve a final cycloheximide concentration of 50 µg/mL, corresponding to a final DMSO concentration of 0.05%. The culture medium in all remaining wells was replaced with this cycloheximide-containing medium. Cells were harvested at 2, 4, 7, 11, and 24 hours after treatment. All cell pellets were collected by centrifugation, washed once with cold PBS, and stored at −80°C until further analysis by Western blotting.

### SDS-PAGE and western blotting

Cell pellets were lysed in RIPA buffer containing 50 mM Tris-HCl pH 7.4, 150 mM NaCl, 1% NP-40, 1 mM EDTA, 1 mM EGTA, 0.1% SDS, 0.5% sodium deoxycholate, and protease/phosphatase inhibitor cocktail (Thermo 78446). We added RIPA buffer at approximately twice the pellet volume and incubated samples on ice for 45 minutes. Lysates were clarified by centrifugation at 16,500×g for 20 minutes at 4°C, and supernatants were collected. Protein concentration was determined using the BCA assay (Thermo 23225).

Samples were prepared in 4× Laemmli buffer supplemented with 10× DTT and heated at 70°C for 5 minutes. We loaded approximately 15 μg of protein per lane onto 10% Bis-Tris gels (1.5 mm, Thermo NP0316BOX) and performed electrophoresis at 200 V for 35 minutes in 1× MES running buffer. Proteins were transferred to PVDF membranes activated in methanol using a wet transfer system at 20 V for 1 hour. Membranes were blocked in 5% non-fat dry milk in TBST (TBS with 0.1% Tween-20) for at least 1 hour at room temperature. Primary antibodies were diluted in TBST containing 1% milk and incubated overnight at 4°C with gentle rocking. The following primary antibodies were used: anti-CLPP (Abcam ab124822, 1:500), anti-ACTB (Abcam ab8224, 1:5000), anti-MALSU1 (Invitrogen PA5-54222, 1:500), anti-MRPS35 (Proteintech 16457-1-AP, 1:5000), anti-MRPL39 (Proteintech 28165-1-AP; 1:500), anti-MRPS28 (Proteintech 16378-1-AP, 1:500), and anti-MRPL28 (Abcam ab126719, 1:500). Following primary antibody incubation, membranes were washed three times for 10 minutes each in TBST and incubated with HRP-conjugated secondary antibodies (anti-rabbit IgG, Cell Signaling 7074; anti-mouse IgG, Cell Signaling 7076; 1:2000 dilution in 1% milk in TBST) for 1 hour at room temperature. After three additional 10-minute washes in TBST, signals were detected using chemiluminescent substrates (SuperSignal West Dura, Thermo 37071; West Pico PLUS, Thermo 34579; or West Atto, Thermo A38554) selected based on signal intensity.

### Oxygen consumption rate

We measured oxygen consumption rate (OCR) using a Seahorse XF96 Extracellular Flux Analyzer (Agilent). HAP1 cells were seeded at 40,000 cells per well in XFe96 Cell Culture Microplates 24 hours prior to measurement. On the day of the assay, culture medium was replaced with XF DMEM medium (pH 7.4, Agilent 103575-100) supplemented with 10 mM glucose, 1 mM pyruvate, and 2 mM L-glutamine. Plates were incubated for 1 hour at 37°C in a non-CO_2_ incubator to allow temperature and pH equilibration. The sensor cartridge was hydrated overnight in XF Calibrant (pH 7.4) at 37°C, and the instrument was calibrated according to manufacturer instructions. OCR measurements were performed using the Mito Stress Test protocol (Agilent 103015-100). We sequentially injected oligomycin (2.0 μM final concentration) to inhibit ATP synthase, FCCP (0.5 μM) to uncouple mitochondrial respiration, and a combination of rotenone and antimycin A (0.5 μM each) to abolish mitochondrial respiration. Following OCR measurements, cells were fixed directly on the assay plate by adding 20 μL of 11% glutaraldehyde (1% final concentration) and incubating for 15 minutes at room temperature on a rocker. Plates were washed three times by decanting into deionized water. Fixed cells were stained with 50 μL of 0.1% crystal violet in 10% ethanol for 15 minutes, then washed extensively with deionized water. After air drying, bound dye was solubilized in 100 μL of 10% acetic acid for 5 minutes at room temperature. Absorbance was measured at 590 nm using a plate reader, and OCR values were normalized to relative cell number determined by crystal violet staining.

### Mitoribosome translation assay

We measured mitochondrial protein synthesis using L-homopropargylglycine (HPG) incorporation followed by click chemistry-based fluorescent detection and flow cytometry, based on modified manufacturer’s instructions (Thermo C10428) and published protocols.^106^ We cultured HEK293T cells (MALSU1-knockout lines generated by CRISPR-Cas9, non-targeting control, MALSU1-overexpressing line, and GFP-overexpressing control) under standard media conditions in 15 cm dishes until reaching 80-90% confluency. We prepared L-HPG at 50 mM (Jena Bioscience, CLK-1067-25), Click-iT HPG reaction buffer and buffer additive (manufacturer’s instructions), methionine-free DMEM (Thermo 21013024) supplemented with GlutaMAX (100×, Thermo 35050-061) and 10% dialyzed FBS, anisomycin at 100 mg/mL in DMSO (Sigma A9789), chloramphenicol at 100 mg/mL in 100% ethanol (Sigma C0378).

Fluorescent dyes were used pre-diluted (Alexa Fluor 488 azide, used for MALSU1-KO condition) or diluted in DMSO for a final concentration at 1 mM (Alexa Fluor 594 azide, used for the MALSU1-OE condition). We reduced culture medium from 25 mL to 10 mL per dish and set up three conditions: no HPG (negative control), HPG with anisomycin (cytosolic translation inhibition only), and HPG with anisomycin plus chloramphenicol (cytosolic and mitochondrial translation inhibition). We added anisomycin to 100 μg/mL final concentration, chloramphenicol to 150 μg/mL. We incubated cells for 30 minutes at 37°C with 5% CO_2_, washed once with pre-warmed PBS, and replaced medium with methionine-free DMEM containing 100 μM HPG, as well as antibiotics according to the three conditions at the same concentrations. We incubated for 1 hour at 37°C with 5% CO_2_. We harvested cells after washing once with PBS and collected them into 15 mL tubes. We resuspended cells in 5 mL mitochondrial protective buffer (10 mM HEPES-KOH pH 7.5, 300 mM sucrose, 10 mM NaCl, 5 mM MgCl_2_) containing 0.0005% digitonin and incubated for 5 minutes at room temperature for selective plasma membrane permeabilization. We pelleted cells at 600×g for 3 minutes, fixed in 1 mL of 4% paraformaldehyde in PBS for 15 minutes at room temperature, then pelleted at 1000×g for 3 minutes. We permeabilized with 1 mL of 0.1% (v/v) Triton X-100 in PBS for 5 minutes, resuspended in 500 μL of 3% BSA in PBS, and pelleted cells again. We performed click chemistry by preparing fresh reaction cocktail per sample (250 μL total): 215 μL 1× Click-iT HPG reaction buffer, 10 μL copper(II) sulfate (4 mM final concentration), 0.625 μL Alexa Fluor 594 azide (1 mM stock) or 0.625 µL Alexa Fluor 488 azide, and 25 μL Click-iT HPG buffer additive (1× final concentration). Pellets were resuspended in this cocktail and incubated for 30 minutes at room temperature protected from light. Washed once with 500 µL Click-iT rinse buffer by centrifugation at 1000×g for 3 minutes, resuspended in 500 μL PBS, and filtered through a cell strainer into FACS tubes. We analyzed samples by flow cytometry using a BD LSRFortessa X-20 (Alexa Fluor 594 azide: laser 561 nm, dichroic filter none, bandpass filter 610/20, PE-TexRed channel; Alexa Fluor 488 azide: laser 488 nm, dichroic filter 505 LP, bandpass filter 530/30, FITC channel), collecting data for at least 10,000 cells per sample with voltages adjusted to maintain signals within dynamic range. We performed gating in FlowJo version 10.10.0 (FSC-A vs SSC-A to exclude debris, SSC-A vs SSC-W for singlets). We calculated geometric mean fluorescence intensity and normalized by subtracting mean background fluorescence from negative controls (no HPG) for each cell line. We ensured equal cell numbers across replicates by randomly sampling to match the lowest count per experimental group. All conditions were performed in three biological replicates.

### siRNA-mediated knockdown

We performed siRNA-mediated knockdown of CLPP in 293T cells using ON-TARGETplus Human CLPP siRNA SMARTpool (Dharmacon L-005811-00-0005) and ON-TARGETplus Non-targeting Pool (Dharmacon D-001810-10-05) as a control. Cells were seeded at 3×10^5^ cells per well in 6-well plates in complete DMEM supplemented with 10% FBS and antibiotics. Twenty-four hours after seeding, cells were transfected using a re-treatment protocol to achieve sustained knockdown. Stock siRNA solutions (20 μM) were diluted to 5 μM in 1× siRNA buffer (Dharmacon B-002000-UB-100). For each well, we prepared two separate tubes in serum-free medium: Tube 1 contained 10 μL of 5 μM siRNA diluted in 190 μL serum-free medium, and Tube 2 contained 4 μL DharmaFECT 1 transfection reagent (Dharmacon T-2001-02) diluted in 196 μL serum-free medium. Both tubes were incubated separately for 5 minutes at room temperature, then combined and incubated for an additional 20 minutes. The siRNA-transfection reagent mixture was then diluted with 1600 μL antibiotic-free complete medium to achieve a final siRNA concentration of 25 nM and 0.2% DharmaFECT 1 in a total volume of 2000 μL per well. To maintain knockdown over extended time periods, we employed a serial passaging and re-treatment strategy. Forty-eight hours after the initial transfection (Day 2), cells were split 1:4 into fresh 6-well plates and re-transfected the same evening using identical transfection conditions. This splitting and re-transfection cycle was repeated on Day 4. Cells were harvested at 48, 96, and 144 hours after the initial transfection, corresponding to timepoints after the first, second, and third transfection rounds, respectively. Samples were analyzed using the proteomics workflow described above. All experiments were performed in triplicate.

### Proteinase K digest

We performed proteinase K digestion to assess the submitochondrial localization of C15orf61-FLAG. We isolated crude mitochondria from HAP1-C15orf61-FLAG cells as described above and resuspended them in isolation buffer (without fatty acid-free BSA or protease inhibitor) at ∼50 μg/μL. We determined protein concentration using a BCA protein assay and diluted samples to 2 μg/μL in isolation buffer without protease inhibitors. We prepared 2× assay solutions (20 μL per condition) on ice using isolation buffer, Proteinase K (NEB P8107S, 0.8 U/μL), 10% digitonin in water (Sigma D141-500MG), and 10% Triton X-100 in water (Sigma X100-100ML): (1) buffer alone (100 μL isolation buffer); (2) PK alone (97 μL isolation buffer, 3 μL PK); (3) PK with 0.01% digitonin (96.8 μL isolation buffer, 3 μL PK, 0.2 μL digitonin); (4) PK with 0.03% digitonin (96.2 μL isolation buffer, 3 μL PK, 0.6 μL digitonin); (5) PK with 0.1% digitonin (95 μL isolation buffer, 3 μL PK, 2 μL digitonin); (6) PK with 0.3% digitonin (91 μL isolation buffer, 3 μL PK, 6 μL digitonin); (7) PK with 1% Triton X-100 (77 μL isolation buffer, 3 μL PK, 20 μL Triton X-100). We aliquoted 20 μL of mitochondrial sample (40 μg protein) into 1.5 mL low-protein-binding tubes on ice using wide-orifice tips. 20 μL of each assay solution was added to designated tubes, flicked 1-3 times, and samples were incubated at 4°C for 30 minutes in a cooling block. We added 2.8 μL of 100 mM PMSF stock to each tube, flicked 1-3 times, then added 16 μL of trichloroacetic acid (TCA; 100% in water) to achieve a final concentration of 25% TCA. We vortexed each sample and incubated on ice for >1 hour. We pelleted precipitated proteins by centrifugation at 16,000×g for 10 minutes at 4°C and removed the supernatant without disturbing the pellet. We washed pellets with 200 μL ice-cold acetone, centrifuged at 16,000×g for 10 minutes at 4°C, removed the supernatant, and dried pellets at ∼85°C for 30 seconds. We resuspended pellets in 20 μL of 1× LDS sample buffer containing 50 mM DTT, vortexed, and heated at 85°C for 10 minutes. We performed electrophoresis and protein transfer according to the western blotting protocol and probed membranes with the following antibodies: Tom20 (Santa Cruz sc-17764, 1:500), Tom22 (CST 90704S, 1:2,000), Timm50 (Proteintech 22229-1-AP, 1:5,000), Timm44 (Proteintech 66149-1-Ig, 1:2,000), HSP60 (EnCor Biotech CPCA-HSP60, 1:5,000), ATPF1A (Proteintech 14676-1-AP, 1:10,000), Diablo (Abcam ab32023, 1:1,000), ATP5ME (Atlas Antibody HPA035010, 1:1,000), and FLAG (Cell Signaling 14793, D6W5B, 1:1,000).

### Mitochondrial membrane potential

We measured mitochondrial membrane potential using flow cytometry to assess mitochondrial function in C15orf61 knockout cells. We cultured HAP1 wild-type, C15orf61-KO1, and -KO2 cells under standard conditions seeded in 15 cm dishes at approximately 80% confluence on the day of the experiment. We harvested cells and pelleted them in 15 mL tubes. We prepared three experimental conditions in triplicate: unstained control, 20 nM TMRM alone, and 20 nM TMRM with 50 μM CCCP for each cell line. We prepared a working solution of 50 μM CCCP by diluting CCCP stock (50 mM in DMSO; Thermo Fisher M20036) 1:1000 in PBS immediately before use. We resuspended unstained and TMRM-only samples in 500 μL PBS and resuspended TMRM+CCCP samples in 500 μL CCCP working solution. We incubated all samples at 37°C for 5 minutes, then added 0.5 μL TMRM stock (20 μM in DMSO; Thermo Fisher M20036) to TMRM and TMRM+CCCP samples for a final concentration of 20 nM. We incubated samples at 37°C for 30 minutes, pelleted cells by centrifugation at 600×g for 3 minutes at room temperature, washed once with 1 mL PBS and resuspended in 1 mL PBS. We kept samples wrapped in aluminum foil at room temperature until analysis. Samples were analyzed on a BD LSRFortessa X-20 flow cytometer using a 561 nm yellow-green laser and 585/15 nm bandpass filter (PE channel) for TMRM detection. We gated for live cells by plotting FSC-A versus SSC-A and then gated for singlets by plotting SSC-A versus SSC-W, using FlowJo version 10.10.0. We collected at least 10,000 events per replicate for all conditions across three biological replicates. We normalized cell counts across replicates by randomly sampling the minimum number of cells observed in any replicate within each condition group.

We calculated geometric mean fluorescence intensity for each sample and normalized values to the mean geometric mean of WT cells stained with TMRM alone.

### ATP synthase activity assay

We measured ATP synthase activity using the MitoCheck Complex V Activity Assay Kit (Cayman Chemical, 701000) following the manufacturer’s protocol with modifications. We isolated mitochondria from cultured cells and resuspended them in Complex V Activity Assay Buffer at 5 mg/mL protein concentration. For each well, we prepared the following reaction mixture: We first added 50 μL of Tube A containing mitochondrial lysate (1 μL mitochondria at 5 mg/mL), Complex V Activity Assay Buffer (48.9 μL), and rotenone (0.1 μL of 1 mM stock in ethanol) to inhibit Complex I activity. We then added 20 μL of either assay buffer alone or assay buffer containing oligomycin (80 μg/mL final concentration, prepared by diluting 10 mM oligomycin stock 1:20 in assay buffer) to determine oligomycin-insensitive background activity. Finally, we initiated the reaction by adding 30 μL of Tube B containing Complex V Assay Enzyme Mix (31.75 μL), Complex V ATP Reagent (1 μL), and Complex V NADH Reagent (1 μL) using a multichannel pipette to ensure simultaneous reaction initiation across wells. We immediately measured absorbance at 340 nm using a microplate reader, recording measurements every 30 seconds for 30 minutes at room temperature. ATP hydrolysis activity was calculated from the linear rate of NADH oxidation (decrease in A340), which is coupled to ATP hydrolysis through pyruvate kinase and lactate dehydrogenase. We calculated specific Complex V activity by subtracting oligomycin-insensitive rates from total rates and normalizing to mitochondrial protein content. All measurements were performed in technical triplicates.

### Cobalamin coenzyme synthesis assay

The synthesis of the different cobalamin coenzymes was determined according to a detailed published method.^107^

### Imaging of mitochondrial network structure

We assessed mitochondrial network morphology by immunofluorescence confocal microscopy. German glass coverslips (12 mm, no. 1½, Fisher 72290-04) were coated with poly-D-lysine (Sigma A-003-E) for 5-10 minutes, washed once with sterile water, and dried under UV light for 1 hour. HAP1 cells were seeded at 5×10^4^ cells per well in 24-well plates containing coated coverslips in IMDM supplemented with glucose, 10% FBS, and penicillin/streptomycin. After overnight culture, cells were fixed with 4% formaldehyde (prepared from 16% stock, Fisher PI28908) in OptiMEM for 5 minutes at room temperature, then washed once with 1× PBS. Fixed cells were permeabilized and blocked in 5% normal goat serum (Fisher 50197Z) in PBS-T (1× PBS with 0.03% Triton X-100) for a minimum of 30 minutes at room temperature. Cells were incubated with primary antibody against HSP60 (Aves Labs CPCA-HSP60, 1:500 dilution in blocking buffer) overnight at 4°C. Following three 10-minute washes in PBS-T, cells were incubated with Alexa Fluor Plus 647-conjugated goat anti-chicken IgY secondary antibody (Invitrogen A32933, 1:500) and DAPI (1:1,000,000 final dilution) for 1 hour at room temperature in the dark. After a final wash in PBS, coverslips were mounted cell-side down on Superfrost Plus microscope slides (Fisher 12-550-15) using Fluoromount-G mounting medium (SouthernBiotech 0100-01, Fisher OB100-01). Excess mounting medium was removed, and coverslips were sealed with clear nail polish (Ted Pella, Fisher NC1849418). Slides were stored sat 4°C in the dark until imaging. Confocal imaging was performed to capture mitochondrial network structure.

### Transmission electron microscopy

HAP1 cells were seeded on coverslips (Electron Microscopy Sciences, 72290-04) using culturing conditions as described above. The next day, media was removed from cells, after which cells were washed with warm 0.15 M cacodylate buffer. Wash buffer was removed and cells were then incubated with fixative (2.5% glutaraldehyde, 2% paraformaldehyde, 0.15 M cacodylate buffer pH 7.4, 2 mM calcium chloride) for 10 min at 37°C. Fixation proceeded overnight at room temperature on an orbital shaker at 300 rpm. Cover slips with cells were processed through infiltration using a Leica EM TP autoprocessor. Cells were rinsed out of fixative with 1X HBSS 3 times for 10 minutes each and subjected to secondary fixation for one hour in 1% osmium tetroxide in 1x HBSS. Following this, samples were rinsed in ultrapure water three times for 10 minutes each and stained for 8 hours in an aqueous solution of 2% uranyl acetate at 4°C. After staining was complete, cells were washed in ultrapure water three times 10 minutes each, dehydrated in a graded acetone series (33%, 66%, 100% x2) for 10 minutes each step, then infiltrated into epon using an Epon 812 (Electron Microscopy Sciences, Hatfield, Pa) resin:acetone series (33%, 66%, 100% for 6 hours each and 2 100% changes 8 hours each). Samples were cured in an oven at 60°C for 72 hours. Post-curing, glass coverslips were dissolved in 42% hydrofluoric acid for 2 hours and the resin embedded cells were mounted for *en face* sectioning. Thin sections 70 nm thick were cut from sample blocks, post-stained with uranyl acetate and Reynolds lead then imaged on a JEOL JEM-1400 Plus (Tokyo, Japan) TEM operated at 120 KeV. Micrographs made using an AMT (Nanosprint15-MkII sCMOS camera).

Quantification was performed by blinded manual tracing of images using the Fiji distribution of the ImageJ software (v2.16.0),^108^ similar as described previously.^109^

### mtDNA quantification by qPCR

Total DNA was extracted from cell pellets using the DNeasy Blood & Tissue Kit (Qiagen 69504) according to the manufacturer’s instructions with minor modifications. Briefly, cells were lysed in buffer ATL containing proteinase K, and 4 μL of RNase A (100 mg/mL, Qiagen 19101) was added prior to addition of buffer AL to eliminate RNA contamination. Following lysis, samples were processed through the DNeasy spin columns, washed, and eluted in buffer AE. DNA concentration was determined by spectrophotometry. Mitochondrial DNA copy number was quantified by duplex TaqMan qPCR using a QuantStudio 6 Flex Real-Time PCR System (Applied Biosystems). We used a FAM-labeled TaqMan assay targeting the nuclear gene B2M (assay ID Hs06637353_s1, catalog 4331182) as an endogenous control and a VIC-labeled, primer-limited TaqMan assay targeting the mitochondrial gene MT-ND1 (assay ID Hs02596873_s1, catalog 4448484) to quantify mtDNA. Each 20 μL reaction contained 10 μL TaqMan Gene Expression Master Mix (2×), 1 μL MT-ND1 assay (20×), 1 μL B2M assay (20×), 1 μL DNA template (40 ng), and 7 μL nuclease-free water. Thermal cycling conditions were: 50°C for 5 minutes, 95°C for 10 minutes, followed by 40 cycles of 95°C for 15 seconds and 60°C for 1 minute. All samples were analyzed in triplicate or quadruplicate. Relative mtDNA copy number was calculated using the ΔΔCt method, with MT-ND1 levels normalized to B2M and expressed relative to control samples.

### Quantitative RT-PCR

We extracted total RNA from cells cultured under standard conditions using the RNeasy Mini Kit (Qiagen, 74104). For cDNA synthesis, we combined 1 μg RNA with 1 μL random hexamer primers (50 ng/μL) and 1 μL 10 mM dNTP mix in a total volume of 10 μL with nuclease-free water. We incubated samples at 65°C for 5 min, then placed them on ice for 1 min. We then added 10 μL cDNA synthesis master mix containing 2 μL 10× RT buffer, 4 μL 25 mM MgCl_2_, 2 μL 0.1 M DTT, 1 μL RNaseOUT, and 1 μL SuperScript III RT (Invitrogen, 18080-051). We performed reverse transcription using the following program: 25°C for 10 min, 50°C for 50 min, and 85°C for 5 min. We diluted cDNA 1:10 in nuclease-free water and used 1.25 μL per qPCR reaction. For qPCR, we prepared 20 μL reactions containing 10 μL PowerUp SYBR Green Master Mix (Thermo Fisher, 4367659), 1 μL primer mix (250 nM final concentration of each primer), 7.75 μL water, and 1.25 μL diluted cDNA template. We performed qPCR using the following cycling conditions: 95°C for 10 min, followed by 40 cycles of 95°C for 15 sec and 60°C for 60 sec, with melt curve analysis. We ran technical triplicates for each sample and normalized gene expression to GAPDH. Primer sequences were: MT-ND3 (forward: CCGCGTCCCTTTCTCCATAA, reverse: AGGGCTCATGGTAGGGGTAA), FASTKD1 (forward: AAGAATTAACTTTTCTGCATTTCCA, reverse: CAGAACAGACACCTCAGTTGGT), GAPDH (forward: TTCGACAGTCAGCCGCATCTTCTT, reverse: GCCCAATACGACCAAATCCGTTGA), and MT-ATP6 (forward: TTCGCTTCATTCATTGCCCC, reverse: GGGTGGTGATTAGTCGGTTGT) as published previously.^47^ Experiment performed twice using four replicates per cell line and target gene.

### Recombinant protein expression and purification

We transformed bacterial expression vectors encoding CBR4 (NM_032783.5) or SDR39U1 (NM_020195.2) into BL21(DE3) cells (Thermo Fisher Scientific, EC0114) and cultured transformants in TB autoinduction medium (BOCA Scientific, GCM19.0500) at 37°C until OD600 reached 0.6. We then shifted cultures to 18°C for 20 hours. We harvested cells by centrifugation, resuspended pellets in lysis buffer (20 mM HEPES, 300 mM NaCl, 10 mM imidazole, pH 7.4), and lysed cells by sonication. We purified CBR4 and SDR39U1 from soluble lysate fractions using HisPur Ni-NTA affinity chromatography (Thermo Fisher Scientific, 25229) with an imidazole gradient (10–200 mM) in 20 mM HEPES, 300 mM NaCl, pH 7.4. We pooled fractions containing purified protein, concentrated using centrifugal filters, and stored aliquots at −70°C until use.

### Differential scanning fluorimetry

Protein melting curves were performed in principle as published previously.^110^ We prepared 20 μL reactions containing 2 μM purified protein, 5× SYPRO Orange dye (Thermo Fisher Scientific, S6650), 100 mM NaCl, and 20 mM HEPES, pH 7.4, in MicroAmp Optical 96-well plates sealed with optical adhesive covers (Applied Biosystems). We measured fluorescence in the ROX channel during a thermal ramp from 25°C to 95°C at 0.015°C/s using an Applied Biosystems QuantStudio 6 Flex Real-Time PCR System. We calculated melting temperatures (Tm) by fitting the transition phase to a two-state Bell’s model.

## Quantification and statistical analysis

### Proteomics data analysis

Data-independent acquisition (DIA) data acquired on the **Orbitrap Astral** mass spectrometer were analyzed using Proteome Discoverer version 3.2.0.450 (Thermo Fisher Scientific). Data processing employed the DIA workflow with the CHIMERYS search engine (run on the Ardia Server). MS1 precursors were used for selection, with isotope pattern reevaluation enabled and profile spectra provided automatically. The Spectrum Selector node applied a signal-to-noise threshold of 1.5 and filtered scans to exclude MS1 spectra. Precursor clipping was set to 2.5 Da before and 5.5 Da after. Searches were conducted against the Homo sapiens UniProt reference proteome using the Inferys 4.7.0 fragmentation prediction model. Trypsin/P was specified as the digestion enzyme with full specificity, allowing up to two missed cleavages, peptide lengths between 7 and 30 amino acids, and charge states of +1 to +4. Carbamidomethylation of cysteine was set as a static modification and methionine oxidation as a dynamic modification, permitting up to three variable sites per peptide. The precursor and fragment mass tolerances were 20 ppm. PSMs were validated using a target-decoy approach with strict and relaxed false discovery rates (FDR) of 1% and 5%, respectively. Quantification was based on MS2 apex intensities (“Quan in all files” mode), storing all feature traces and the best PSM per peptide.

Consensus workflow analysis was performed to integrate and validate peptide and protein identifications. PSM grouping, peptide validation, and protein filtering were applied sequentially, retaining only peptides of high confidence (minimum length six amino acids). Protein-level validation was conducted with strict and relaxed FDR thresholds of 1% and 5%. Quantification was performed using unique and razor peptides, considering protein groups for peptide uniqueness. Protein abundances were calculated as the summed peptide intensities and normalized to the total peptide amount per sample. Scaling was disabled, and no data imputation or hypothesis testing was applied within Proteome Discoverer.

Data-dependent acquisition (DDA) data acquired on the **Orbitrap Exploris 240** mass spectrometer were analyzed using Proteome Discoverer version 2.5.0.400 (Thermo Fisher Scientific). The analysis comprised a processing workflow for peptide identification and quantification and a consensus workflow for validation, alignment, and protein-level roll-up. Spectra were processed using the Sequest HT search engine with Percolator validation. MS1 precursor-based quantification was performed with CID fragmentation. The Minora Feature Detector was used for chromatographic feature detection (minimum trace length 5, S/N ≥ 1, ΔRT ≤ 0.2 min), linking only high-confidence peptide-spectrum matches (PSMs). Spectra were filtered to include full-scan MS2 events with precursor masses between 350–5000 Da and an FT signal-to-noise threshold of 1.5. Searches were performed against a UniProt human proteome FASTA database using trypsin specificity (max 2 missed cleavages), precursor and fragment mass tolerances of 10 ppm and 0.6 Da, respectively, and a minimum peptide length of 6 amino acids. Static carbamidomethylation (+57.021 Da, C) and dynamic oxidation (+15.995 Da, M), N-terminal acetylation (+42.011 Da), methionine loss (−131.040 Da), and methionine loss + acetylation (−89.030 Da) were included. Up to four variable modifications and three equal modifications per peptide were allowed. Spectral matches were filtered by Percolator using a concatenated target-decoy approach with q-value–based validation at strict and relaxed false discovery rates (FDR) of 1% and 5%, respectively. In the consensus workflow feature alignment and precursor-based quantification were performed using the Feature Mapper and Precursor Ions Quantifier nodes. Retention times were aligned across runs (maximum shift 10 min, 10 ppm mass tolerance), and precursor abundances were determined from MS1 intensities.

Quantification was based on unique + razor peptides, normalized by total peptide amount, and scaled across all samples. Protein abundances were calculated as summed peptide intensities, and ratios were derived by pairwise comparison with background-based t-testing. PSM grouping and peptide validation used automatic q-value control at 1% (strict) and 5% (relaxed) FDR. Only high-confidence peptides (minimum length 6) were retained. Protein-level validation was set to 1% (strict) and 5% (relaxed) FDR, and annotation included peptide flanking residues, modification sites, and positions within master proteins.

Quantified protein intensity matrices were processed in Perseus software version 4.1.3.0 for data normalization and missing value imputation.^111^ Protein abundances were log_2_-transformed prior to imputation. Missing values were imputed separately for each sample from a normal distribution simulating signals from low-abundance proteins, using a downshift of 1.8 and a width of 0.3 relative to the observed data distribution. For analysis of endogenous XL-AEMS data, this downshift was increased to 3-3.5 when a protein was not detected in any replicates across all conditions but one. Statistical analysis was performed in R version 4.3.1, where log_2_ fold changes between sample groups and corresponding p-values were calculated using a two-sided Welch’s t-test. Processed proteomics data is provided in **Table S11**.

### Mitochondrial gene sets

Mitochondrial gene sets were defined based on the MitoCarta3.0 inventory and the corresponding MitoPathways3.0 annotations,^17^ with several modifications to improve coverage and annotation consistency. ATP synthase gene symbols were updated to match official nomenclature. The mitoribosome category was expanded to include distinct subgroups representing the small (mtSSU) and large (mtLSU) subunits. A separate category was introduced for mtDNA-encoded genes. Within the Coenzyme Q_10_ metabolism pathway, additional subcategories were created to distinguish proteins associated with Complex Q and proteins involved in the synthesis and attachment of the isoprenoid tail. The complete list of curated mitochondrial gene sets used in this study is provided in **Table S12**.

### Transcript-protein decoupling metric

We quantified transcript–protein decoupling by computing residuals from within-KO linear regressions. For each KO we fit an ordinary least-squares model regressing log_2_ fold change proteomics on log_2_ fold change transcriptomics: log_2_fc_prot = β_0_ + β_1_·log_2_fc_tx + ε. From each fitted model we extracted both raw residuals (εlJ) and studentized residuals (standardized by their estimated standard errors) using standard R diagnostic functions. Each gene was assigned a category label (mitochondrial genes in the MitoCarta3.0 reference set versus the remainder). To evaluate decoupling at the gene set level we mapped genes to mitochondrial gene programs (according to MitoCarta3.0) and retained only programs represented by at least n_min genes (default n_min = 6). For each program we computed three dispersion statistics on the standardized residuals: standard deviation (SD), interquartile range (IQR), and median absolute deviation (MAD). Programs with high dispersion indicate that protein abundances systematically deviate from the transcript-level prediction within that KO, reflecting post-transcriptional or translational regulation. We visualized program-level decoupling by plotting dispersion (y-axis, log_10_ scale) against programs ordered by that metric (x-axis), with point size encoding the number of genes per program; selected programs of biological interest were color-coded and labeled. All computations used grouped tidy data operations, and MAD was chosen as the primary spread metric for robust quantification of decoupling magnitude across programs.

### Principal component metric

We characterized the dominant axis of transcript–protein covariation for each gene using principal component analysis. For every gene we extracted all KO:gene rows excluding self-targeting perturbations and performed PCA on the bivariate distribution (log_2_fc transcriptomics, log_2_fc proteomics) without scaling (scale = FALSE) to preserve the native variance contribution of each modality. From each gene-specific PCA model we extracted the first principal component loading vector (PC1) and computed two summary statistics: the proportion of variance explained by PC1 (var_explained = sdev_1^2^ / sum(sdev^2^) · 100) and the orientation angle of PC1 with respect to the transcriptomics axis (angle = atan2(PC1_y, PC1_x) · (180 / pi), in degrees). The angle quantifies the relative weighting of transcript versus protein changes along the dominant axis of variation: angles near 0 degrees indicate transcript-driven variation, angles near 45 degrees indicate coordinated transcript–protein changes, and angles near 90 degrees indicate protein-driven variation decoupled from transcripts. To facilitate interpretation, we transformed negative angles by adding 180 degrees, so all angles fall in the range 0 to 180 degrees, with an additional transformation to center angles near 180 degrees on the left side of the x-axis for visualization continuity. For gene set-level analysis we assigned genes to functional categories (according to MitoCarta3.0 pathway definitions), expanded the table to include all matching categories per gene, and filtered to the biologically interpretable range (angle_positive2 between −10 and 100 degrees). Pathway-specific distributions were visualized as scatterplots of angle versus variance explained, with reference lines at 45 degrees (balanced regulation) and 90 degrees (protein-dominated regulation), and background density from all genes for context.

### Nearest neighbor distance analysis

We computed nearest-neighbor distances to detect KO:gene outliers in the joint space of effect size (log_2_ fold change) and statistical significance. For each KO:gene row we first protected p-values by replacing non-positive or missing p with a tiny floor epsilon (1e-300) and defined the significance score S = -log10(max(p, epsilon)). Within each gene we then scaled effect sizes and significance to produce coordinates (x, y) by x = effect / max_KO(|effect|) and y = S / max_KO(S); rows for which a gene has zero or undefined maxima were left with missing coordinates. All NN computations were performed within each gene: for the set of finite (x, y) points we used a fast k-NN search (Euclidean metric) with a bounded heuristic k (default k = 10) and, if no valid neighbor was found among the first k, a fallback exhaustive search with k = n (all other points). From each ordered neighbor list we accepted the nearest candidate only if it satisfied three biological filters: different KO (exclude self and same-KO replicates), same sign of effect (sign(effect_nbr) = sign(effect_ref)), and smaller magnitude (|effect_nbr| < |effect_ref|); if no neighbor passed these filters the NN distance was recorded as missing. Distances were computed as d_ij = sqrt((x_i - x_j)^2^ + (y_i - y_j)^2^). To make distances comparable across genes and modalities (transcriptomics, proteomics) we normalized all non-missing raw NN distances by the global median of the NN table for that modality: d_norm = d / median(d_non-missing), and used log(1 + d_norm) for visualization. We recorded missingness as frac_NA = number(d = NA) / N_total (proteomics = 0.99%, transcriptomics = 0.99%). For combined analyses we removed any row missing either transcript or protein normalized NN distance (1.97% of all possible NN distances excluded). Implementation used a high-performance k-NN routine (nn2), standard tidy data tooling for grouping and filtering, and analytical plots of (log(1 + d_norm,tx), log(1 + d_norm,prot)) with optional thresholds (example thresholds: t_prot = 2, t_tx = 2).

### Entropy calculation

We quantified gene-level variability across KO perturbations using Shannon entropy as a measure of distributional spread. For each mitochondrial gene (MitoCarta3.0 reference set) we collected on log_2_ fold change values a gene-by-gene basis across all KOs, excluding the KO targeting that gene itself to avoid confounding self-targeting effects. We binned each gene’s log_2_fc distribution into n_bins equally spaced intervals (default n_bins = 20) spanning the observed range [min(log_2_fc), max(log_2_fc)], computed bin frequencies f_i, normalized to probabilities p_i = f_i / Σf_i, and calculated Shannon entropy as H = -Σ p_i · log(p_i + ε), where ε = 10^-10^ prevents numerical issues when bins are empty. High entropy indicates dynamic behavior across perturbations; low entropy indicates static behavior. This procedure was applied independently to transcriptomics and proteomics. To classify genes we computed the mean (μ_H) and standard deviation (σ_H) of entropy across all mitochondrial genes within each modality and defined three categories: “Static” (H < μ_H - k·σ_H), “Intermediate” (μ_H - k·σ_H ≤ H ≤ μ_H + k·σ_H), and “Dynamic” (H > μ_H + k·σ_H), where k is a tunable threshold factor (default k = 1). Classification was performed separately for transcriptomics and proteomics to account for modality-specific variability.

### Embedding of mitochondrial genes

We embedded mitochondrial genes in low-dimensional space to visualize KO-specific perturbation patterns. For each modality (transcriptomics, proteomics) we first computed a composite distance metric that integrates effect magnitude and statistical confidence. Within each KO we converted raw p-values to q-values using the Storey method and defined a signed distance measure: d = (1 - q) · (2/(1 + 2^(-log_2_fc)) - 1), where the first term weights by significance and the second term is a sigmoid-transformed effect size that saturates at ±1 for large fold changes. Data were restructured into wide format with genes as rows and KOs as columns (each cell containing d for that gene-KO pair) and filtered to retain only mitochondrial genes from the MitoCarta3.0 reference set. We computed uniform manifold approximation and projection (UMAP) embeddings using the cosine distance metric with modality-specific tuning: transcriptomics used n_neighbors = 8, spread = 11; proteomics used n_neighbors = 10, spread = 0.3. All embeddings projected to two components (n_components = 2) with a fixed random seed (seed = 42) for reproducibility. Point density in the resulting UMAP space was calculated using a nearest-neighbor kernel method: for each gene we counted neighbors within a radius r^2^ = (Δx_range + Δy_range)/70 (where Δx_range and Δy_range are the data ranges along UMAP1 and UMAP2), weighting distances anisotropically to account for axis-scale differences, and normalized densities to [0,1] by dividing by the maximum. Density visualization used log_2_-transformed counts. The identical pipeline was applied independently to transcriptomics and proteomics data, yielding separate embeddings for comparative analysis.

### Analysis of complexome profiling data

#### Wild-type sample processing and normalization

We processed complexome profiling data from three biological replicates of HAP1 wild-type cells. We imported protein abundance data from Proteome Discoverer output files and tidied the data by truncating gene symbols at semicolons to remove isoform annotations. For duplicate gene symbols, we retained the entry with the highest number of unique peptides, and if tied, the highest number of PSMs. We replaced non-predominant duplicate gene symbols with their accession numbers, filled missing gene symbols with accession numbers, removed proteins absent from all fractions, and replaced remaining NA values with zero. We normalized protein abundances in two sequential steps. First, we normalized each protein’s abundance within each gel fraction by dividing by the total protein abundance in that fraction. Second, we normalized each protein across all fractions by dividing by its maximum abundance value, scaling all values to 0-1. We estimated molecular weight for each gel fraction using an exponential decay model fitted to protein markers (NativeMark Unstained Protein Standard, Invitrogen LC0725). We fitted a linear model on log-transformed molecular weights and predicted values for all 60 fractions using MW = A × exp(B × slice). We retained only proteins detected in all three biological replicates for downstream analysis and merged data into a single table containing gene names, sample identifiers, replicate numbers, estimated molecular weights, and normalized abundances.

#### Correlation and UMAP analysis

We performed correlation analysis to identify proteins with similar migration patterns. We used the normalized data and calculated Pearson correlation coefficients for all unique protein pairs. For complex V subunit analysis, we calculated mean Pearson correlation coefficients between each protein and all complex V subunits. We performed UMAP analysis to visualize protein clustering based on migration profiles. We filtered data to include only mitochondrial proteins, removed proteins absent from all fractions, and converted to wide format with proteins as rows and gel fractions as columns. We applied the umap function in R using n_neighbors = 20, n_components = 2, spread = 8, and metric = “cosine”, with random seed 42 for reproducibility. We annotated UMAP coordinates with protein complex membership using mitochondrial pathway annotations.

#### Individual replicate analysis and visualization

We analyzed individual replicates separately following the same data tidying, normalization, and molecular weight estimation procedures. We generated heatmaps for specific protein complexes using hierarchical clustering (Euclidean distance, complete linkage) to order genes. For oxidative phosphorylation complex visualization, we calculated complex-level abundances by averaging normalized abundances of all subunits within each complex for each gel fraction, then normalized by dividing by the maximum value for each complex. We visualized complex migration patterns as heatmaps and line-density plots, *e.g.*, we created a vertical heatmap showing all three biological replicates side-by-side for each OxPhos complex (complex I-V) to visualize reproducibility across replicates.

### Analysis of external complexome profiling datasets

We compared C15orf61 migration patterns with OxPhos complexes across multiple independent complexome profiling datasets. We imported previously processed and normalized complexome profiling data from three studies via the CEDAR platform. From CRX42^75^ we used high-resolution clear native gel electrophoresis (hrCNE) data from untreated control HEK cells, averaging technical replicates. From CRX47, we used control HEK cell samples with averaged data across replicates. From CRX38^76^ we used control HEK cells (Control_01) processed by high-resolution clear native PAGE. We merged wild-type control samples from each study, retaining gene names, normalized relative abundances, gel slice numbers, study identifiers, and estimated molecular weights in kDa. We filtered this dataset for OxPhos protein sets and calculated mean relative abundance for each protein complex at each molecular weight across gel slices. We assigned C15orf61 as its own category and labeled complex I-V subunits according to their respective complex and visualized as density plots.

### mtDNA association analysis

We analyzed associations between mtDNA levels and gene expression using linear regression modeling. We used log_2_ fold changes of mtDNA quantity as well as transcripts and proteins to performed two complementary linear regression tests. First, to identify genes whose expression levels predict mtDNA quantity, we modeled log_2_(median mtDNA ratio) as the outcome and gene log_2_ fold changes as predictors. Second, to identify genes whose expression is predicted by mtDNA levels, we reversed the model structure using lm(log_2_fc_gene ∼ log_2_(mtDNA)). For each gene, we extracted the regression coefficient (beta, effect size) and p-value from model summaries. We performed these analyses separately for transcript and protein data, then merged results by gene symbol to create a combined table containing effect sizes and p-values for both transcripts and proteins in both modeling directions.

### DepMap CRISPR screen data analysis

We analyzed the impact of C15orf61 and complex V knockouts on cell fitness using publicly available CRISPR screening data from the Cancer Dependency Map. The DepMap Public 25Q2+Score, Chronos; downloaded on 16 May 2024). We imported gene effect scores quantifying the impact of gene knockout on cell growth across cancer cell lines, where negative scores indicate growth inhibition upon knockout and positive scores indicate growth enhancement upon knockout. We extracted C15orf61 gene effect scores and calculated mean gene effect scores for complex V subunit genes for each of the 1,183 cell lines. The data was visualized as individual data points per cell line as well as summarized as density plot.

### Quantification of confocal images

We quantified mitochondrial network connectivity from confocal microscopy images using automated image analysis. We imported .czi microscopy files into ImageJ and applied an ICA (Independent Component Analysis) filter followed by auto contrast adjustment using a custom macro, then exported processed images as .tiff files. We analyzed mitochondrial network architecture using Nellie, an automated organelle analysis platform that segments and quantifies morphological features without requiring manual calibration or parameter optimization.^112^ Nellie processed the .tiff images and generated output .csv files containing measurements at the organelle level and branch level. We calculated mitochondrial network connectivity as the ratio of total branches to total organelles by dividing the number of rows in the branches output file by the number of rows in the organelle output file. Higher branch-to-organelle ratios indicate more connected, reticulated mitochondrial networks, while lower ratios indicate more fragmented networks. We also quantified total branch length per organelle from the organelle-level output file as an additional metric of network complexity.

### Integrated mtDNA related phenotypes

To systematically assess relationships between mitochondrial genome regulation, gene expression, and cellular phenotypes across knockout cell lines, we performed dimensionality reduction using UMAP. We compiled five metrics for each knockout line: (1) mtDNA quantity measured by ddPCR (log_2_-transformed ND1:B2M ratios), (2) mtDNA transcript levels (mean log_2_ fold-changes of mtDNA-encoded genes from RNA-seq), (3) mtDNA-encoded protein levels (mean log_2_ fold-changes from proteomics), (4) mitochondrial mass (mean log_2_ fold-changes of significantly altered mitochondrial proteins, p < 0.05), and (5) cell growth (log_2_ fold-changes in cell density). We z-score normalized each metric across all cell lines to enable comparable weighting in the embedding. The following parameters were used for UMAP calculation: n_neighbors = 5, min_dist = 0.1, metric = “euclidean”, n_components = 2, random_state = 123, with spectral initialization. This generated a two-dimensional embedding where proximity reflects similarity across the integrated phenotypic space. We visualized each metric independently using color gradients (blue for decreased, red for increased values) to identify knockout cell lines exhibiting coordinated changes across multiple molecular and cellular layers.

### Analysis of Perturb-Seq data

Genome-scale Perturb-seq data from K562 cells were obtained from the Genome-wide Perturb-seq resource^27^ and processed datasets were downloaded from Figshare (https://plus.figshare.com/articles/dataset/20029387) on 9 December 2023. For analysis of specific knockdown targets (*e.g.*, MMADHC) we extracted normalized bulk expression data and ranked all detected transcripts by z-score.

### Analysis of mito-nuclear balance data

We analyzed published data from a genome-wide CRISPR screen assessing mito-nuclear protein balance.^78^ We extracted gene effect scores (Combo casTLE Effect) and confidence scores (Combo casTLE Score) from the supplementary dataset (sheet casTLE_GateD from Supplementary Table 2). We inverted the gene effect scores (×-1) for intuitive interpretation, such that positive values represent increased ratios of mtDNA-encoded COX1 to nuclear-encoded COX4 upon gene knockout. Confidence scores reflect the statistical reliability of each measurement. We removed duplicate gene entries, retaining unique symbols for analysis. We visualized knockout targets ranked by confidence score, with color gradient indicating the direction and magnitude of the effect on COX1:COX4 ratio.

### Additional resources

An interactive platform for our data will be available upon peer-review.

## Data Availability

All data produced in the present study are available upon reasonable request to the authors.

## ACKNOWLEDGMENTS

We thank the Pagliarini Lab for their helpful feedback and discussion throughout the duration of this study, Sergej Djuranovic and Slavica Pavlovic-Djuranovic for technical advice and sharing of equipment regarding mitoribosome sucrose sedimentation, Jarred Rensvold for technical assistance, Robi Mitra for guidance on RNA-Seq data collection and analysis, Richard Rodenburg for sharing information of *MALSU1* patient variants, Robert Taylor for assistance with GeneMatcher inquiries, D. Sean Froese and Vito R.T. Zanotelli for sharing *MMADHC* patient cell lines and biochemical data, Matthew Stefely for visual graphics support, the Washington University Diabetes Research Center (NIH 5P30 DK020579) for use of instrumentation, and the Washington University Center for Cellular Imaging (WUCCI; supported by Washington University School of Medicine, The Children’s Discovery Institute of University and St. Louis Children’s Hospital (CDI-CORE-2015-505 and CDI-CORE-2019-813) and the Foundation for Barnes-Jewish Hospital (3770 and 4642)). This work was supported by the European Molecular Biology Organization (ALTF 263-2022) and the Swiss National Science Foundation (P500PB_211038) (both to P.F.), NIH training grants T32GM140935 and T32AG000213 (both to A.Y.S.), NIH award R35 GM131795 (to D.J.P.), and funds from the BJC Investigator Program (to D.J.P.). D.J.P. is an investigator of the Howard Hughes Medical Institute. This article is subject to HHMI’s Open Access to Publications policy. HHMI lab heads have previously granted a nonexclusive CC BY 4.0 license to the public and a sublicensable license to HHMI in their research articles. Pursuant to those licenses, the author-accepted manuscript of this article can be made freely available under a CC BY 4.0 license immediately upon publication.

## Author contributions

Conceptualization: P.F., D.J.P. Data curation: P.F. Formal analysis: P.F., A.Y.S., D.J.P. Funding acquisition: P.F., D.J.P. Investigation: P.F., M.F., A.J.S., K.L. Methodology: P.F., M.F. Project administration: P.F. Resources: D.J.P. Software: P.F., A.Y.S. Supervision: P.F., D.J.P. Validation: P.F., M.F. Visualization: P.F. Writing – original draft: P.F., D.J.P. Writing – review & editing: P.F., D.J.P. All authors critically reviewed and approved the final version of the manuscript.

## Declaration of AI-assisted technologies in the writing process

During the preparation of this work the authors used the Claude and ChatGPT platforms to assist in proof reading and light editing. After using this service, the authors reviewed and edited the content as needed and take full responsibility for the content of the published article.

## SUPPLEMENTAL INFORMATION

All data produced in the present study are available upon reasonable request to the authors.

- **Table S1**. Standardized residuals from linear regression comparing transcript and protein levels across cell lines organized into mitochondrial gene sets, related to **Fig. 2**.
- **Table S2**. Nearest neighbor distances, related to **Fig. 2**.
- **Table S3**. Principal component 1 and entropy metrics, related to **Fig. 4, 5**.
- **Table S4**. Processed complexome profiling data of WT, ATPAF2-, ATP5ME-KO cells, related to **Fig. 5**.
- **Table S5**. Processed complexome profiling data of WT, C15orf61-KO1/2 cells, related to Fig. 5.
- **Table S6**. Processed complexome profiling data of GFP-, C15orf61-OE cells, related to Fig. S6.
- **Table S7**. mtDNA quantification data and linear regression calculations, related to **Fig. 6**.
- **Table S8**. Processed RNA-seq data table, related to STAR Methods.
- **Table S9**. Updated HAP1 KO cell collection proteomics data table, related to STAR Methods.
- **Table S10**. Merged transcriptomics and proteomics table with processed data, related to STAR Methods.
- **Table S11**. Processed proteomics data from experiments performed in this study, related to STAR Methods and **Fig. 2, 3, 4, 5, 6, S2, S4, S6, S7**.
- **Table S12**. Customized mitochondrial gene sets based on MitoCarta3.0, related to STAR Methods.

## DATA AND CODE AVAILABILITY

All data produced in the present study are available upon reasonable request to the authors.

- Raw RNA sequencing data are deposited on the ArrayExpress platform under accession number E-MTAB-16262.
- Raw mass spectra files from proteomics experiments performed in this study are deposited on the MassIVE platform under project number MSV000100243.
- Processed proteomics data are provided as Supplemental Information (**Table S11**).
- Analyses and data can be explored online upon peer-review.
- Additional information needed to reanalyze the data in this paper can be obtained by contacting the lead author.

## Supplementary Figures

**Figure S1.**
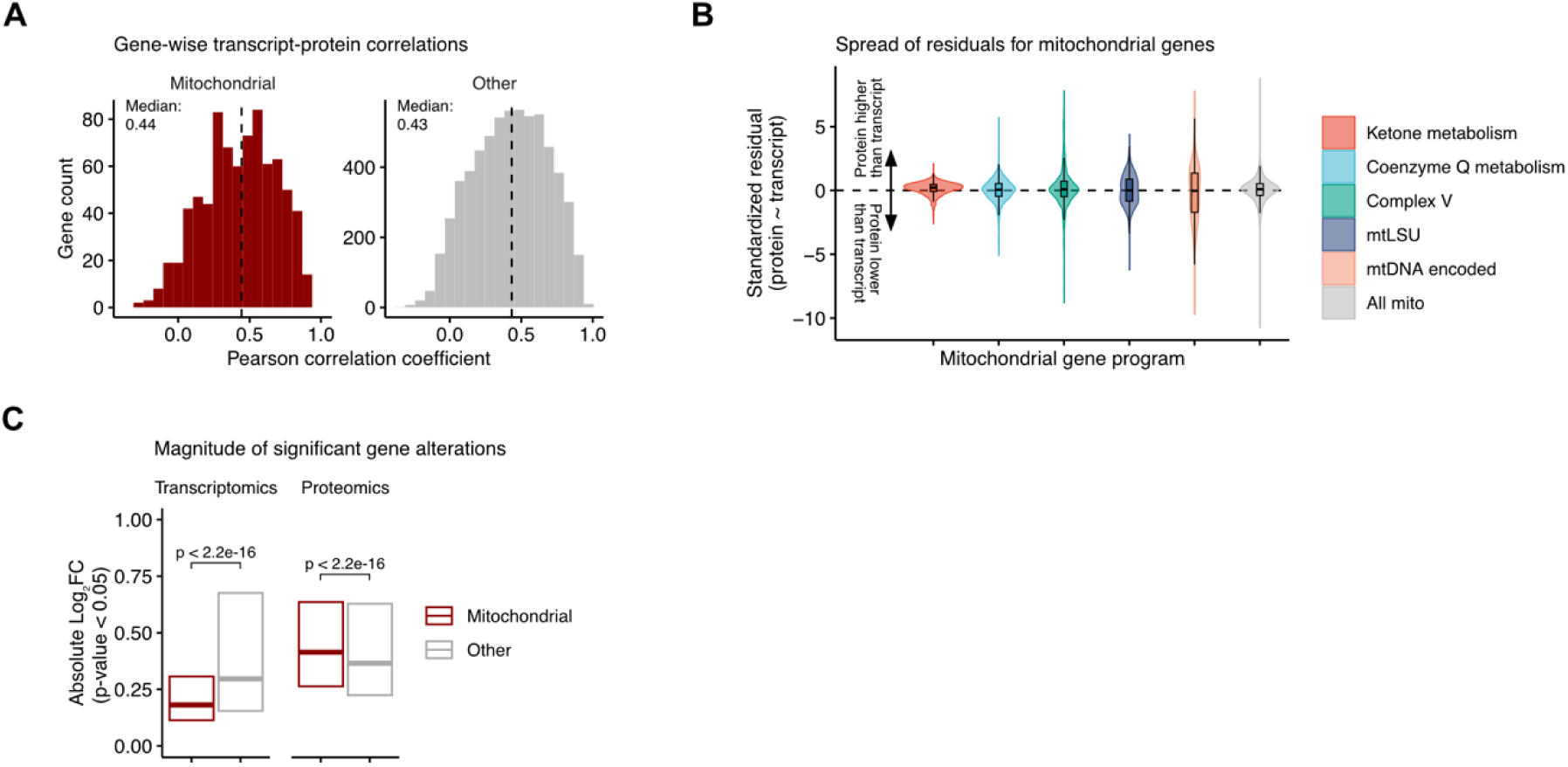
General transcript-protein comparison metrics. (A) Histogram of Pearson correlation coefficients calculated on a gene-by-gene basis, split into mitochondrial and non-mitochondrial genes. (B) Standardized residuals for each gene were extracted from linear regression models and then summarized into mitochondrial gene sets, of which five examples are highlighted here compared the overall spread of all mitochondrial genes. Box plots show median, interquartile range (IQR), and whiskers extending to 1.5×IQR. (C) Box plots comparing absolute log_2_ fold changes in transcriptomics and proteomics data, that met a p-value cut-off of 0.05, split into mitochondrial and non-mitochondrial genes. Boxes indicate median and 25^th^, and 75^th^ percentile, respectively. Statistical comparison is based on Wilcoxon rank-sum test.

**Figure S2.**
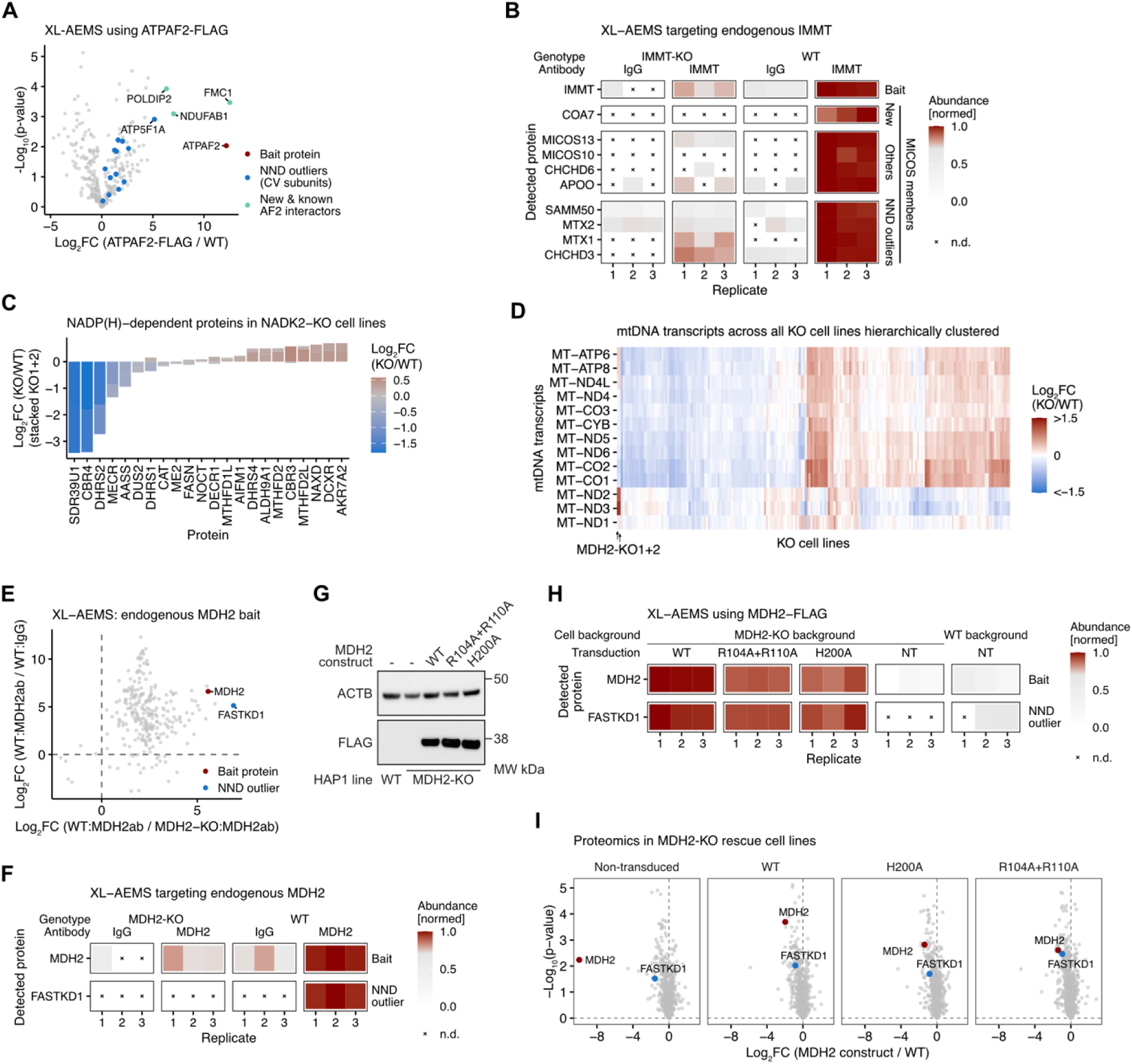
Validation experiments for protein-protein relationships predicted by the nearest neighbor distance analysis. (A) Volcano plot of cross-linking affinity enrichment mass spectrometry (XL-AEMS) experiment performed using a FLAG-tagged ATPAF2 protein as bait. (B) Selected proteins from the XL-AEMS experiment using IMMT and IgG control antibodies. Abundance values were z-transformed. (C) Proteomics data derived from NADK2-KO replicates clones for known NADP(H)-dependent proteins (defined by UniProt and manual curation). Bar plots are stacked log_2_ fold change values derived from two different knockout clones. (D) Heatmap of all mtDNA-encoded transcripts detected in our dataset across all 203 knockout cell clones. The MDH2-KO clones are highlighted and show an exclusive elevation of MT-ND3. The color gradient is capped at an absolute log_2_ fold change of 1.5. (E) Scatter plot of XL-AEMS experiment using an MDH2 antibody to target the endogenous protein in wild-type mitochondria from HAP1 cells. (F) Selected proteins from the XL-AEMS experiment using MDH2 and IgG control antibodies. Abundance values were z-transformed. (G) Western blot in wild-type and MDH2-KO, rescued with three different FLAG-tagged MDH2 constructs. (H) Selected proteins from the XL-AEMS experiment using three different FLAG-tagged MDH2 constructs. Abundance values were z-transformed. (I) Volcano plots based on proteomics data obtained from HAP1 MDH2 rescue cell lines, compared to wild-type.

**Figure S3.**
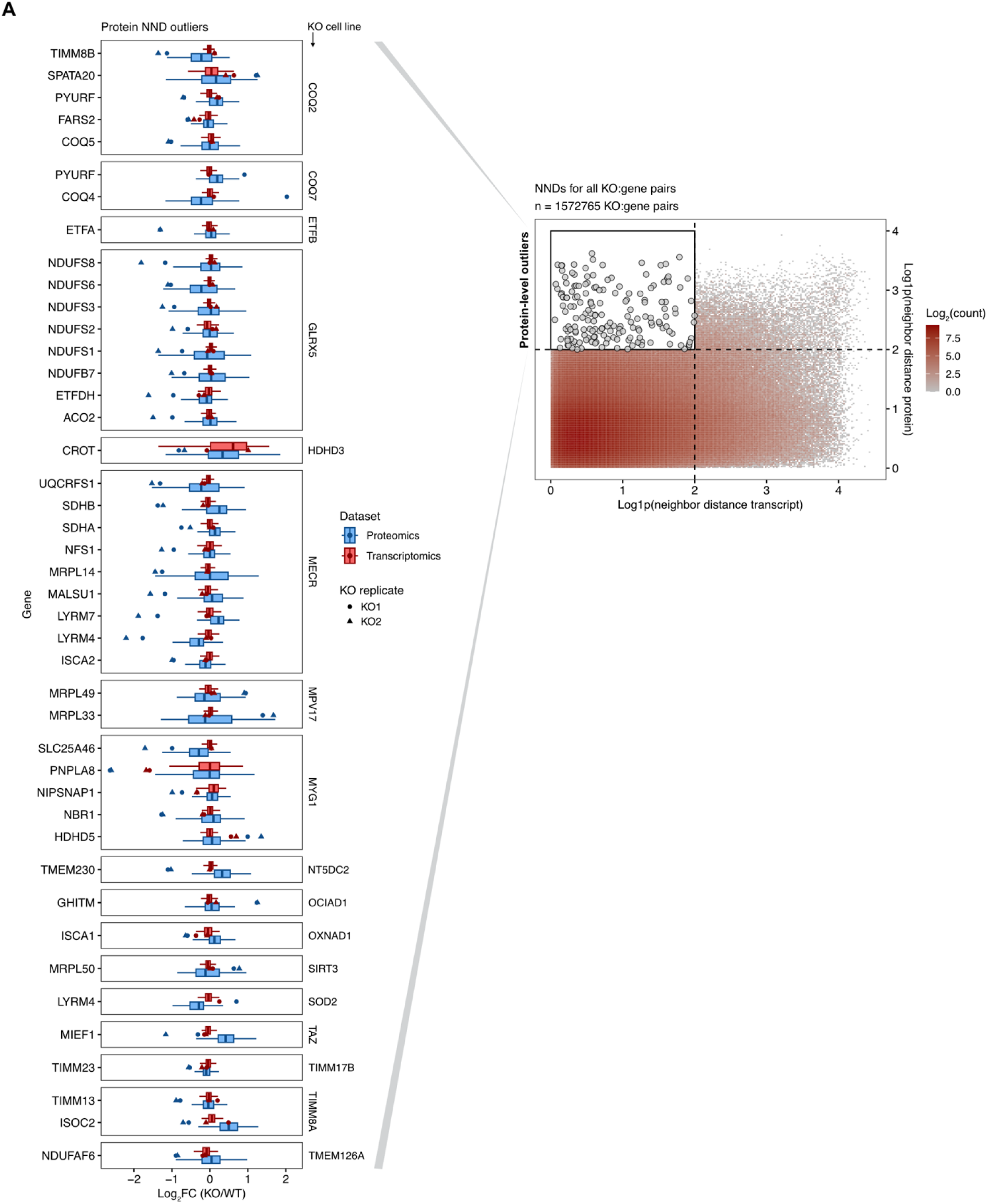
Nearest neighbor distance analysis protein-level outliers. (A) Compiled hits of KO-gene:protein relationships predicted by the nearest neighbor distance outlier analysis, based on the following thresholds: log1p(transcript) < 2, log1p(protein) > 2, absolute log2 fold change protein > 0.5. A few examples just below those cut-offs were added for specific interest (COQ2, COQ7, NT5DC2, SOD2, and TAZ).

**Figure S4.**
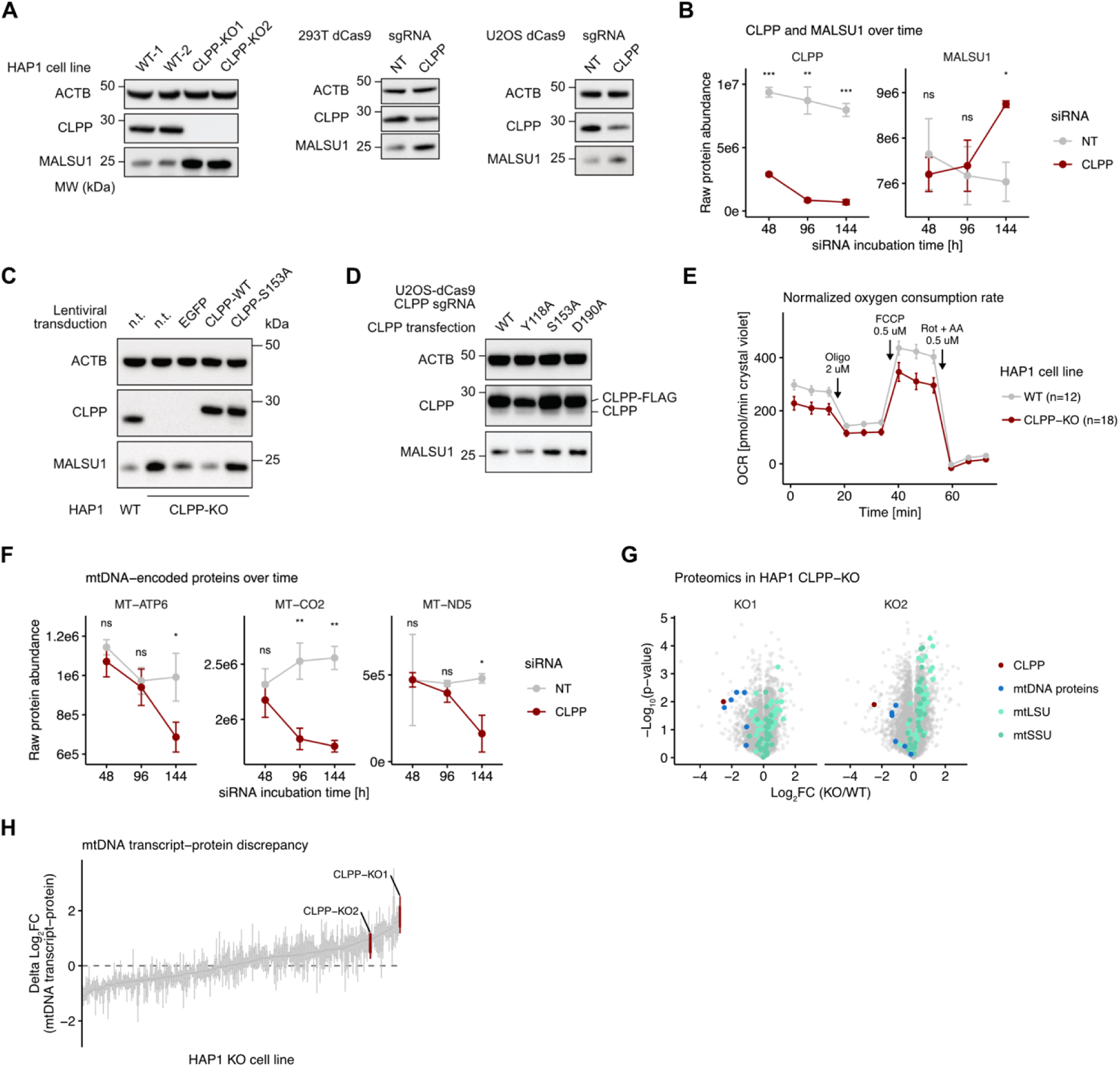
Validation of MALSU1 as CLPP substrate and phenotypic effects of CLPP-KO and MALSU1 modulation. (A) Western blot targeting CLPP and MALSU1 in three different cellular systems with absent or reduced CLPP protein levels. (B) Raw protein abundance values of CLPP and MALSU1 as assessed by mass spectrometry using cells transfected with siRNA (NT, non-targeting; CLPP, targeting CLPP) and incubated for three different time points. Data is based on three biological replicate measurements for each time point; error bars indicate standard deviation; statistical significance was assessed using Welch’s t-test; p-values ≤0.05 (*), ≤0.01 (**), ≤0.001(***). (C) Western blot for MALSU1 and CLPP performed using lysates from HAP1 CLPP-KO cells stably transduced with different CLPP constructs. (D) Western blot for MALSU1 and CLPP performed using lysates from U2OS-CRISPRi-CLPP cells transfected with different CLPP constructs (Y118A, constitutively active; S153A and D190A, inactive). (E) Oxygen consumption rate assessed using a Seahorse assay and different mitochondrial inhibitors. Replicates represent individually platted cells in different wells. (F) Same siRNA experiment as above for three mtDNA-encoded proteins. Statistical significance was assessed using Welch’s t-test; p-values ≤0.05 (*), ≤0.01 (**). (G) Volcano plots of proteomics data from HAP1 CLPP-KO cell clones. Different proteins and protein groups of interest are highlighted. (H) Box plots of differentials between transcript and protein levels of all mtDNA-encoded genes per HAP1 KO cell line. CLPP-KO replicate clones are highlighted.

**Figure S5.**
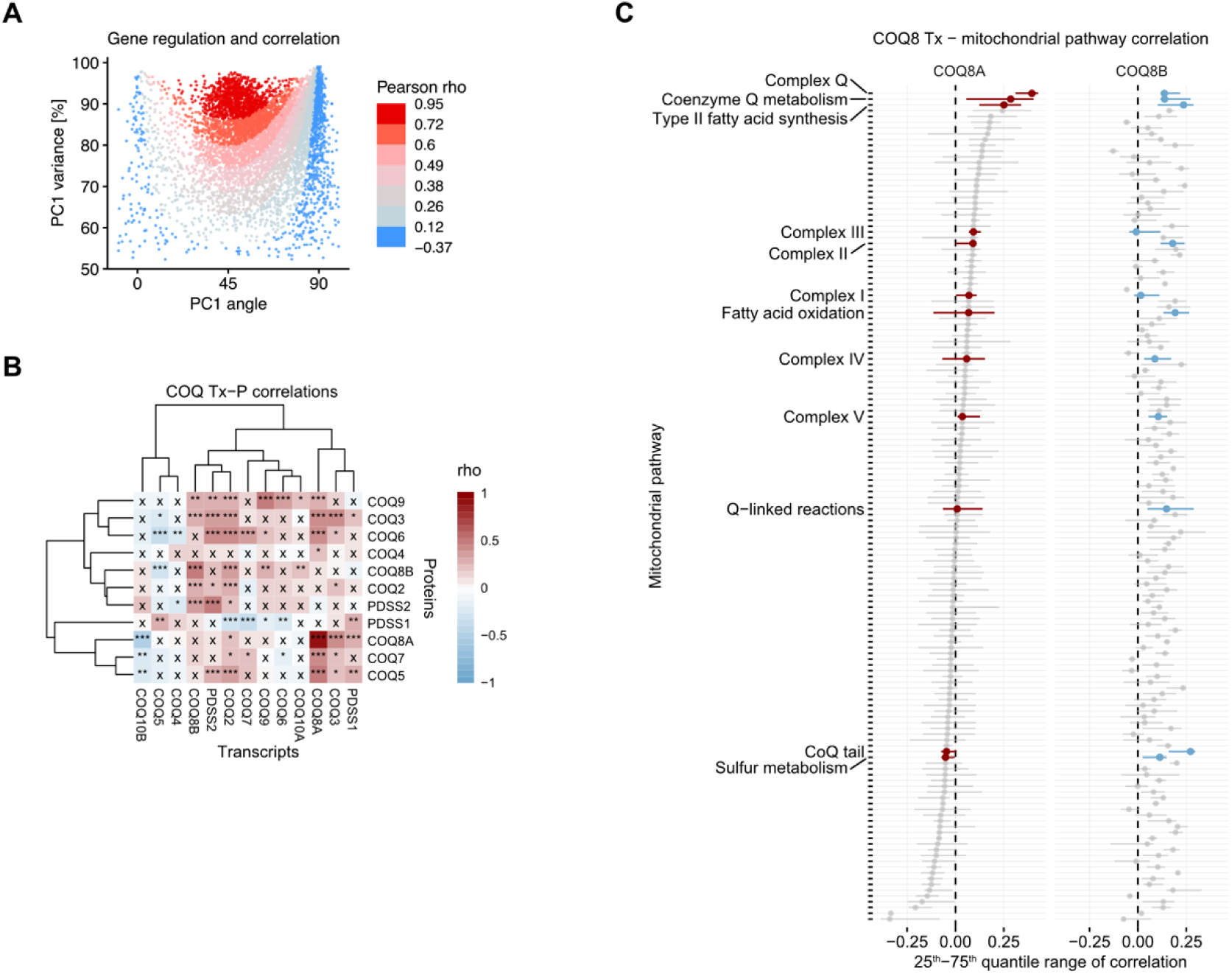
Transcript-protein relationships with a focus on the CoQ biosynthetic pathway. (A) Same molecular regulation metric plot based on principal component analysis as presented in main Fig. 4. Pearson correlation coefficients were calculated for each gene’s transcript-protein relationship across knockout cell lines and is indicated according to the color code. (B) Heatmap depicting Spearman correlation coefficients of CoQ gene transcript-protein comparisons. Asterisks indicate p-values >0.01 (x), <0.01 (*), <0.001 (**), <0.0001(***). (C) Spearman correlation between COQ8A/COQ8B transcript and mitochondrial protein levels across knockout conditions. Point range plot shows the distribution of correlation coefficients between each COQ8 paralog and gene sets, with pathways ordered by median correlation strength.

**Figure S6.**
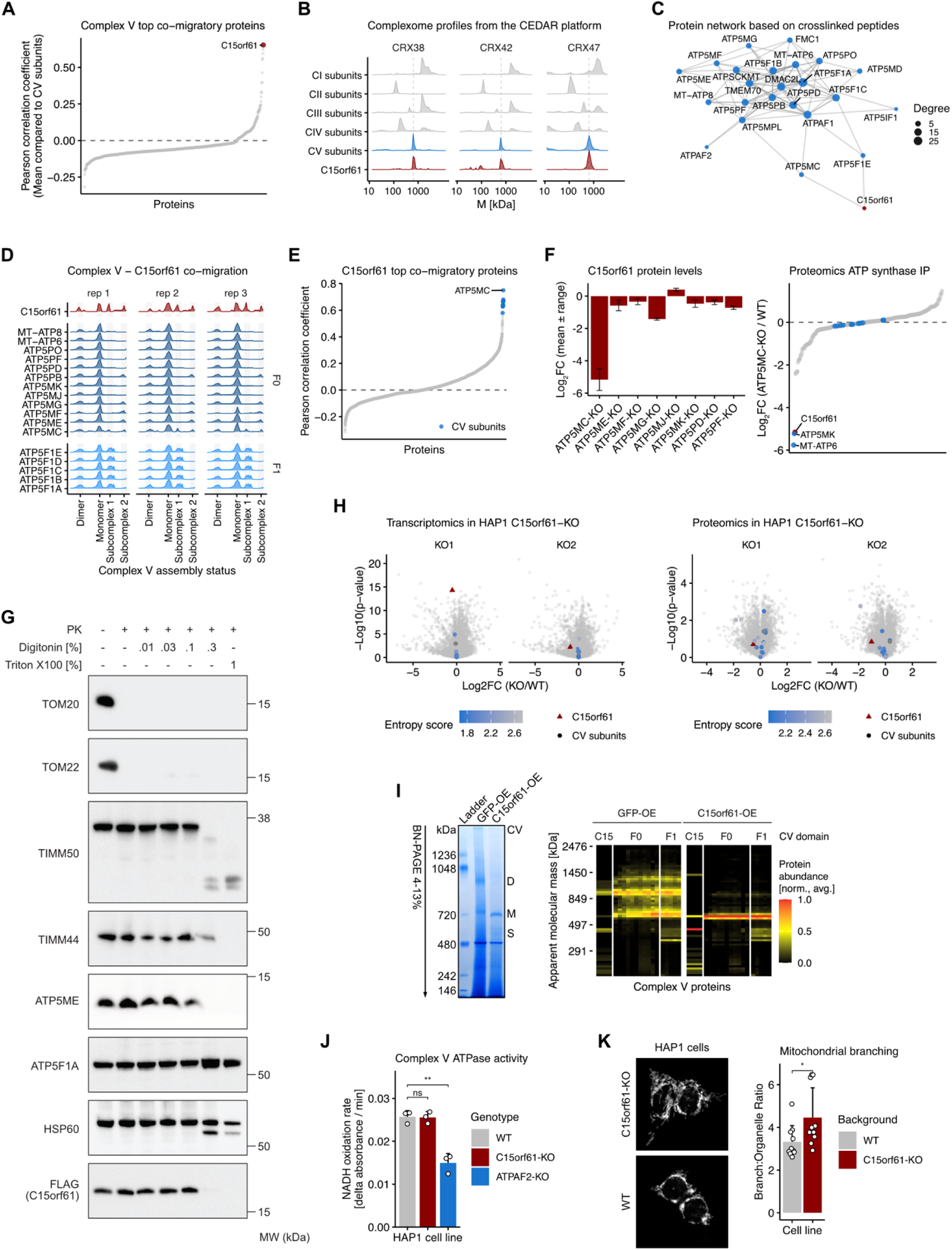
C15orf61 interaction validation, complexome profiling, and further characterization. (A) Rank plot of complexome migration profile correlation coefficients (each protein compared to all CV subunits, averaged). (B) Analysis of three public complexome profiling datasets obtained from the CEDAR platform.^115^ Average of OxPhos complex subunits and C15orf61 profiles are shown as density plots. (C) Network plot based on crosslinking peptides identified following XL-AEMS experiment using C15orf61-FLAG as bait protein. (D) Individual migration profiles of all CV subunits and C15orf61 across all three wild-type complexome profiling replicates. (E) Rank plot of complexome migration profile correlation coefficients (C15orf61 compared to each protein). (F) Left: C15orf61 protein levels in multiple public proteomics datasets.^74,116,117^ Error bars indicate range of two individual replicates. The ATP5MC is a triple knockout of all three c-ring genes (ATP5MC1-3). Right: Proteomics data based on ATP synthase precipitation from HAP1 ATP5MC-KO cells. (G) Proteinase K assay probing for multiple mitochondrial outer and inner membrane, and matrix proteins. (H) Transcriptomics and proteomics data in HAP1 C15orf61-KO replicate clones. CV subunits are highlighted and overlayed with the entropy/variability score. (I) Complexome profiling of mitochondria extracted from HAP1 with either GFP or C15orf61 overexpression. Heatmaps depict individual CV proteins in columns. (J) Bar plot depicting ATPase activity of complex V using mitochondrial isolated from three different HAP1 cell lines. Statistical significance was determined using Welch’s t-test. Statistical significance was assessed using Welch’s t-test; p-values ≤0.01 (**). (K) Confocal images of HAP1 wild-type and C15orf61-KO cells using HSP60 staining as a mitochondrial marker. Quantification metric is branches per organelle as a proxy for mitochondrial network connectivity. Statistical significance was assessed using Welch’s t-test; p-value ≤0.05 (*).

**Figure S7.**
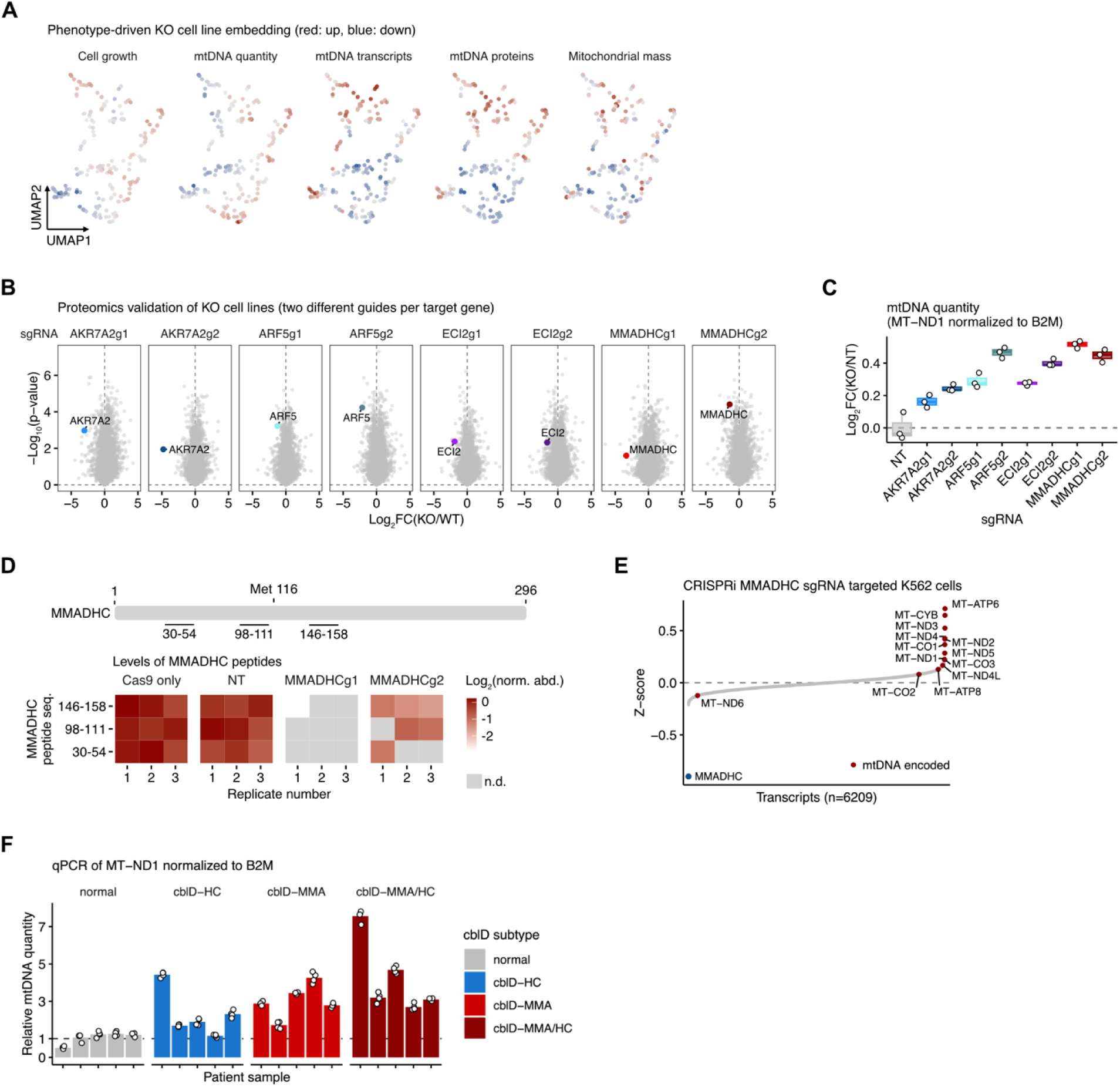
mtDNA related phenotypes and MMADHC validation experiments. (A) UMAP embedding of HAP1 knockout cell lines based on integrated phenotypic measurements (mtDNA quantity, mtDNA transcripts and proteins, mitochondrial mass, and cell growth), colored by z-scored metric values (red: increased, blue: decreased). (B) Volcano plots based on proteomics experiments in CRISPR/Cas9-KO cell lines. Each gene was targeted with two different guides, generating two biological replicate clones. (C) mtDNA quantification using qPCR using DNA extracts derived from CRISPR/Cas9-KO cell lines. (D) Proteomics validation of 293T CRISPR/Cas9-MMADHC-KO cell lines at the peptide level, indicating that peptides across both N- and C-terminal domains are strongly reduced. (E) Published Perturb-seq data^27^ indicating a strong increase of mtDNA-encoded transcripts in MMADHC knockdown cells. (F) Individual mtDNA measurements of patient fibroblast lines, each assessed in technical quadruplicates.

